# The resilient brain against childhood poverty

**DOI:** 10.1101/2024.06.18.24308965

**Authors:** Bowen Hu, Tongxi Yang, Yuanyuan Hu, Shaozheng Qin

## Abstract

Poverty remains a persistent structural issue in society, profoundly impacting children’s mental health and resilience. However, the influence of poverty on children’s resilience and its neural underpinnings is not well understood. This study investigates this impact using a nationally representative sample (N = 11878) and neuroimaging techniques to explore the heterogeneity of resilience mechanisms in poor children. Our findings reveal that poor children (living below the federal poverty threshold) experience exacerbated effects of early life adversities (ELAs) on behavioral problems, which persist into adolescence, indicating disrupted resilience. By subtyping poor children based on neural representations of self-regulation (brain activation during stop signal task), we identified two distinct subtypes: subtype-1, with heightened neural activation, exhibited significantly worse resilience; while subtype-2, with reduced activation, showed resilience levels comparable to non-poor children. Two subtypes did not differ significantly in superficial characteristics including ELA exposure and demographics. However, poor children with subtype-1 had thicker left-middle-frontal-gyrus (L-MFG) brain regions, correlating with fewer behavioral problems and weaker ELA impacts, suggesting a unique resilience mechanism. This L-MFG-based resilience mechanism is exclusive to subtype-1, and not observed in subtype-2 or non-poor children. These findings underscore the importance of understanding individual heterogeneity in resilience mechanisms among disadvantaged children. Our study emphasizes the need for future research to delve deeper into adaptation processes to ELAs and poverty, advocating for the exploration of varied resilience mechanisms to mitigate the stigma associated with poverty and guide interventions aimed at narrowing mental health gradient.

## Introduction

67.4% children around the world (Salmeron Gomez et al., 2023) and 16.3 % children in the U.S. (Shrider & Creamer, 2023) live in poverty. Growing up in poverty can have detrimental effects on children’s overall well-being, with these negative impacts persisting into adolescence and developing into lifelong consequences (Brooks-Gunn & Duncan, 1997; Victora et al., 2022). Among these consequences, the influence of childhood poverty on mental health stands out as a widely researched topic (Udalova et al., 2022). This is attributed to the fact that compromised mental health not only can exacerbates/drive/foster/entrench intergenerational poverty through various pathways (Harper et al., 2003; C. A. McEwen & McEwen, 2017; McLeod & Shanahan, 1993; Tribble & Kim, 2019; Victora et al., 2022), but also carries profound neural underpinnings that impact various dimensions of well-being (B. S. McEwen & Gianaros, 2010; C. A. McEwen & McEwen, 2017; Monk & Hardi, 2023; Rakesh et al., 2023; Tooley et al., 2021). Although listed as the top priority among the United Nations’ Sustainable Development Goals, eradicating childhood poverty presents hard challenges. Nevertheless, understanding the mechanisms underlying the impact of childhood poverty may help us strive to mitigate its impact.

The impact of childhood poverty on mental health has been attributed to more exposure to early life adversities (C. A. McEwen & McEwen, 2017; Tooley et al., 2021; Tribble & Kim, 2019). Children from low-income families experience a higher incidence of ELAs, including family turmoil, violence, noise, etc. (Evans, 2004; C. A. McEwen & McEwen, 2017; Rakesh et al., 2023), the cumulative effect of which poses a significant risk factor for mental health (Daníelsdóttir et al., 2024). While emphasizing the impact of exogenous risk factors (i.e., ELAs) on mental health is reasonable, it is equally important not to overlook the endogenous resilience in children. Robust resilience empowers children with the capacity to adapt and buffer the influences from adverse environmental influences (Holz et al., 2020), thereby maintaining relatively positive psychological outcomes following adversity exposure (Rutter, 2006). Moreover, given the inevitable high levels of adversity exposure experienced by poor children, the effective (rather than disrupted) functioning of resilience is indispensable for their psychological well-being.

However, consensus regarding the manifestation of resilience in poor children remains elusive. On one hand, excessive exposure to ELAs could disrupt resilience through allostatic overload (B. S. McEwen et al., 2015). Meanwhile, childhood poverty often signifies the absence of promotive and protective factors and processes, such as prosocial involvement and parental support, which are essential components of resilience (C. A. McEwen & McEwen, 2017; Rutter, 2012; Ungar & Theron, 2020). On the other hand, the resilience framework highlights the role of adaptation, conceptualizing the experience of adversity as a form of practice where heightened exposure may fosters plasticity and adaption in the stress-response system (Colich et al., 2020; Nederhof & Schmidt, 2012; Rutter, 2006; Snell-Rood & Snell-Rood, 2020). Research also indicates that among children who have experienced high levels of poverty-related ELAs, there are still some children, particularly those with high levels of self-regulation, who exhibit relatively good mental health (De France et al., 2022; Ehrlich et al., 2024) academic performance (Brody et al., 2013, 2020). Furthermore, the inconsistency between research evidence and theoretical perspectives suggests the presence of individual heterogeneity, suggesting that the impact of childhood poverty on resilience may vary among poor children.

Self-regulation and its neural substrates are pivotal in fostering resilience, particularly among poor children (Brody et al., 2013; De France et al., 2022; Flouri et al., 2014). First, self-regulation capacity and its neural representations are central to mental health (Robson et al., 2020) and resilience against adversities (Brody et al., 2013; Ehrlich et al., 2024; Kalisch et al., 2015; C. A. McEwen & McEwen, 2017; Ungar & Theron, 2020). Additionally, childhood represents a critical period for the development and neurodevelopment of self-regulation (McClelland & Cameron, 2012; Pas et al., 2021; Raffaelli et al., 2005), during which divergent development trajectories emerge (Montroy et al., 2016). Specifically, during the transition from childhood to adolescence (i.e., in late childhood), poor children lacking parental caregiving may experience unmet safety needs, which could either ripen their self-regulation capacity for independent safety provision (Rakesh et al., 2023), or lead to gradual self-regulation depletion due to elevated allostatic load (Evans & Kim, 2013). In conclusion, the heterogeneity of neurodevelopment in self-regulation may significantly influence the development of either robust or weak resilience in poor children.

In this study, by leveraging the heterogeneity of neural representations of self-regulation, we aim to distinguish between robust and weak resilience among poor children facing ELAs. Considering the widespread and significant impact of childhood poverty, and aiming to ensure that research outcomes can effectively inform pertinent policies and services nationwide, we examined data from a substantial population-based cohort of 9-10-year-old children across the United States, from Adolescent Brain Cognitive Development (ABCD) Study (Garavan et al., 2018). The neural representations of self-regulation are obtained through modeling the brain activation during the stop signal task (Logan, 1994), a well-established assessment tool for sensitively evaluating the neural activity and neurodevelopment associated with children’s self-regulation function (Pas et al., 2021). Then, the subgroups of poor children characterized by heterogeneous neural representations of self-regulation was derived by a multivariate semi-supervised machine learning algorithm (Heterogeneity through Discriminative Analysis, HYDRA). Finally, we examined whether non-poor children and the subgroups of poor children presents difference in terms of resilience and other profiles, during childhood (baseline) and adolescence (three waves of follow-up). Robust resilience is defined as the ability to withstand the impact of ELAs with minimal effects on mental health, operationalized as a less profound effect (i.e., weaker impact strength) of (cumulative) ELAs on mental health (Kalisch et al., 2015).

## Methods

### Sample

The Adolescent Brain Cognitive Development Study recruited 11878 children (aged 9 to 10 years at baseline) from 22 sites in the U.S., representing the broad U.S. child population (Garavan et al., 2018). Only children proficient in English, free from medical, neurological, or cognitive issues, , and without contraindications for MRI were included (Garavan et al., 2018). The ABCD Study obtained approval from the local institutional review board (IRB) at 22 data collection sites. Parents provided written informed consent at each site, while youth participants provided assent (Thompson et al., 2019). Detailed information regarding ABCD project can be found at https://abcdstudy.org/about/.

### Measures

The neuroimaging data, self-report demographic data (including race, sex, family income, and number of people living in the home), and data on early life adversities (ELAs) were collected at baseline. Behavior problem data were collected at baseline and three subsequent follow-up waves (at approximately one-year intervals).

### Poverty

We computed the income-to-needs ratio for each child’s family based on parent reports of annual family income and the number of people living in the household, utilizing the federal poverty threshold as a reference (Health & Services, 2017). The income-to-needs ratio was computed as the family income divided by the applicable federal poverty threshold. Children from families with an income-to-needs ratio below 100% were classified as poor children.

### Early life adversity (ELAs) and cumulative ELAs

We analyzed 14 types of ELAs across various domains including individual, family, school, and community (refer to Figure 1A or Table X for a list of specific variables; see Supplementary Material S1 for detailed descriptions of their measurement). These ELAs encompassed variables such as experience of trauma, family conflict, bad environment in school, neighborhood unsafety, and, others. The ELAs comprised both categorical variables (e.g., experience of trauma) and continuous variables (e.g., family conflict), wherein categorical variables were coded as 0 (no exposure) or 1 (exposure), and scores for continuous variables were scaled to a range from 0 (least severe exposure) to 1 (most severe exposure) for subsequent calculation of cumulative ELAs using a cumulative score.

**Figure 1.**
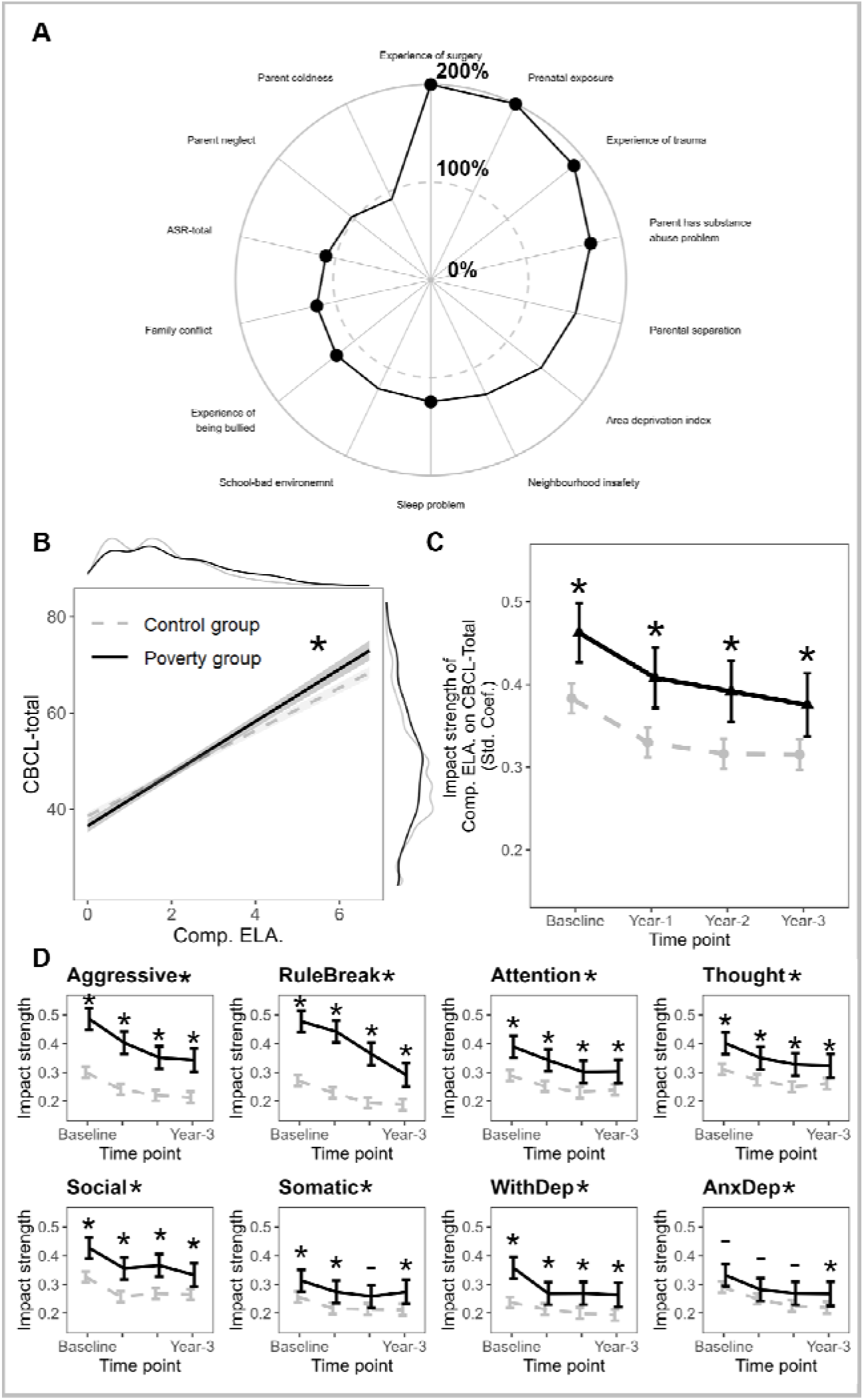
Modulation of cumulative ELAs’ impact on behavior problems by childhood poverty. By including the interaction term between group (poverty vs. non-poverty) and (cumulative) ELAs in the linear mixed effect models to predict CBCL-total score, we evaluated whether the impact of (cumulative) ELAs on CBCL-total score varied across groups. **A,** ratio of standardized coefficients for various ELA types of the poor children to non-poor children. Dots represent interaction significance (p < 0.05). **B,** differences in cumulative ELAs’ impact between groups, with solid and dashed lines denoting poor and non-poor children, respectively. Shaded areas indicate 95% confidence intervals (CIs), and variable distributions are depicted by density plots. **C and D**, trends of the impact strength of baseline-measured cumulative ELAs on CBCL-total score (C) and CBCL subscale scores (D) over time between groups, with error bars indicating the 95% CIs. Abbreviation: ELAs, early life adversities.

### Behavior problems

Children’s behavior problems were assessed at each wave using the Child Behavior Check-List (CBCL), a 113-item questionnaire completed by parents to evaluate dimensional psychopathological syndromes in children and adolescents (Achenbach, 2001). Parents rated a series of behavior problems observed in their children over the past six months using a 3-point scale (0 = Not True; 1 = Somewhat/Sometimes True; 2 = Very True/Often True). The scores for all items were summed to calculate a total score. Additionally, we calculated scores for 8 specific subscales, including aggressive behavior, anxious/depressed, attention problems, rule-breaking behavior, somatic complaints, social problems, stress problems, thought problems, and withdrawn/depressed. These subscales were used to capture more nuanced differences in behavior problems. We used norm-referenced T-scores of CBCL for all the analysis.

### Neuroimaging

We utilized tabulated ROI-based data from the ABCD Data Release 5.0 as the primary source of neuroimaging data for our study, which included data from the stop-signal task fMRI scans and structural MRI scans. These datasets underwent rigorous quality control and preprocessing by the ABCD study group (Casey et al., 2018; Hagler et al., 2019; see Supplementary Material S3 for details). Cortical areas were delineated using the Destrieux atlas (Destrieux et al., 2010), comprising 148 ROIs, while subcortical areas were delineated using the ASEG Atlas (Fischl et al., 2002, p. 20), comprising 30 ROIs.

For the analysis of stop-signal task fMRI data, we modeled a contrast between correct stop and correct go conditions to derive activation maps related to the process of inhibitory control (Chaarani et al., 2021; van Rooij et al., 2015). From these maps, we extracted beta weights from each ROI to serve as a neural representation of self-regulation. Additionally, for the analysis of structural MRI data, we measured thickness in each cortical ROI and volume in each cortical and subcortical ROI.

### Cognitive function

Apart from all the variables mentioned above, we also examined the difference between groups in cognitive function. Cognitive function of the children was measured by seven tasks in NIH toolbox (Casaletto et al., 2015; Thompson et al., 2019; Weintraub et al., 2014). These seven tasks assessed different dimensions of cognitive function, including language vocabulary knowledge, inhibitory control, etc. (see Supplementary Material S2 for details). We used norm-referenced T-scores of the seven tasks and their composite scores (crystallized cognition score, fluid cognition score, and total score) for all the analysis. Details regarding the instruments can be found in Supplementary Material S2.

### Statistical Analyses

The threshold for statistical significance was set at *p* <.05, multiple comparisons (between types of ELAs, waves, subscales, and their combinations) were adjusted with FDR method.

### Missing data

We excluded all participants with missing demographic data and behavior problem data at baseline. Given the significant exclusion rate (up to XX%) of participants due to quality control measures applied to neuroimaging data, we retained these participants in analyses not involving neuroimaging data. Prior to computing cumulative ELAs, missing data for each type of ELA were imputed with corresponding mean values.

### Identifying heterogeneous subtypes of neural representations

The Heterogeneity Through Discriminative Analysis (HYDRA) algorithm was employed to identify heterogeneous subtypes of neural representations of self-regulation. HYDRA was a non-linear multivariate semi-supervised machine learning algorithm designed to derive unique biomarkers for heterogeneous subtypes of clinical samples, integrating binary classification and subtype clustering (Varol et al., 2017). In brief, the HYDRA algorithm utilizes a support vector machine framework with k (user-specified) linear hyperplanes to classify clinical and control samples. Each individual within the clinical sample is optimally discriminated from the control samples on a specific hyperplane, thereby defining the subtype to which the clinical sample belongs. In this study, we used the neural representation of self-regulation (beta weights across 178 ROIs) as features, and used poverty or non-poverty as labels for clinical or control samples. For detailed modeling procedures and determination of the optimal number of subtypes, please refer to Supplementary Material S4.

### Comparison of resilience between groups

In this study, one of our primary aims is to compare resilience between non-poor children and poor children, as well as between non-poor children and two subtypes of poor children. Robust resilience is operationally defined as a weaker impact of (cumulative) ELAs on behavior problems. To examine this, behavior problems were treated as the response variable, and the interaction terms between (cumulative) ELAs and group categories were included as predictors in our analysis. This approach allowed us to assess whether the impact strength of (cumulative) ELAs varied significantly across different groups or whether this impact was moderated by group membership.

We employed linear mixed-effects models using the ’lme4’ package in R (Douglas Bates et al., 2015) to model these associations. We controlled for fixed effects including age, sex, race/ethnicity, and income-to-needs ratio, as well as random effects for site and family nested within site. Furthermore, when extending our analysis to data from four waves, we included the three-way interaction term between cumulative ELAs and groups and time points as a predictor, and included subject ID as an additional random effect.

### Comparison of other profiles between groups

In this study, we also aimed to compare various profiles, including demographic characteristics, exposure (levels) to ELAs, beta weights of brain activation, cortical thickness, etc., between different groups. These comparisons were conducted using linear mixed-effects models, controlling for the same set of confounding variables as in the previous step. However, in specific cases, additional confounding variables were controlled for. When analyzing models that included beta weights of brain activation (as in Figure 2A), we additionally controlled for the fixed effects of head motion (mean framewise displacement in mm). Similarly, when analyzing models that included cortical thickness (either as a dependent variable as in Figure 3C, or as an independent variable as in Figure 3D), we additionally controlled for the fixed effects of mean cortical thickness across all ROIs.

**Figure 2.**
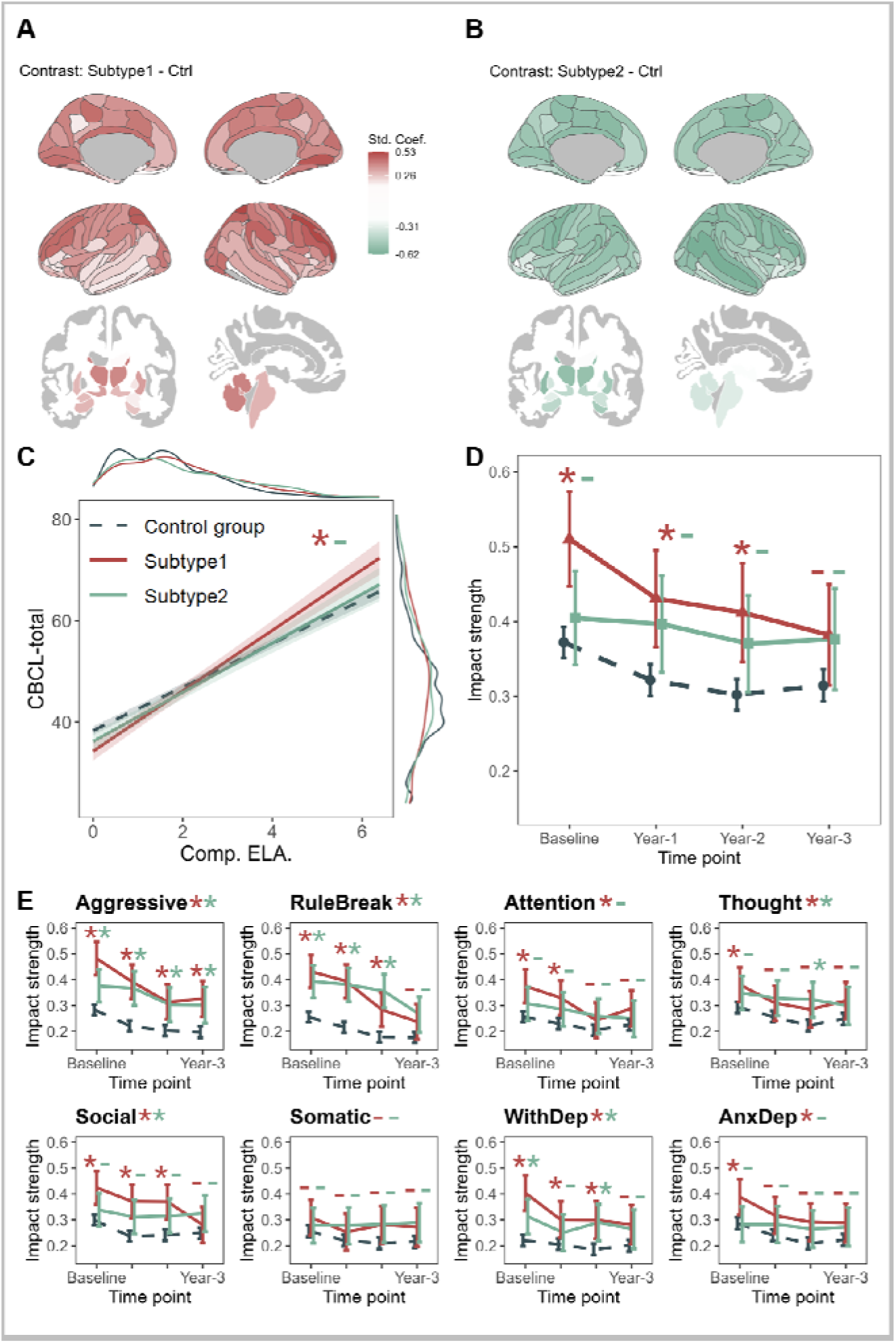
Modulation of cumulative ELAs’ impact on behavior problems by neuroimaging phenotypes among poor children. **A and B,** regional differences among non-poor children and poor children with heterogeneous neuroimaging phenotype in brain activation during the stop signal task. **C,** differences in cumulative ELAs’ impact among groups, with solid line representing poor children (different neuroimaging phenotypes shown in red and green), and dashed line indicating non-poor children. Shaded areas represent the 95% CIs, and variable distributions are visualized by density plots on the edges. **D and E**, trends of the impact strength of baseline-measured cumulative ELAs on CBCL-total score and CBCL subscale scores over time between groups, with error bars indicating the 95% CI.

**Figure 3.**
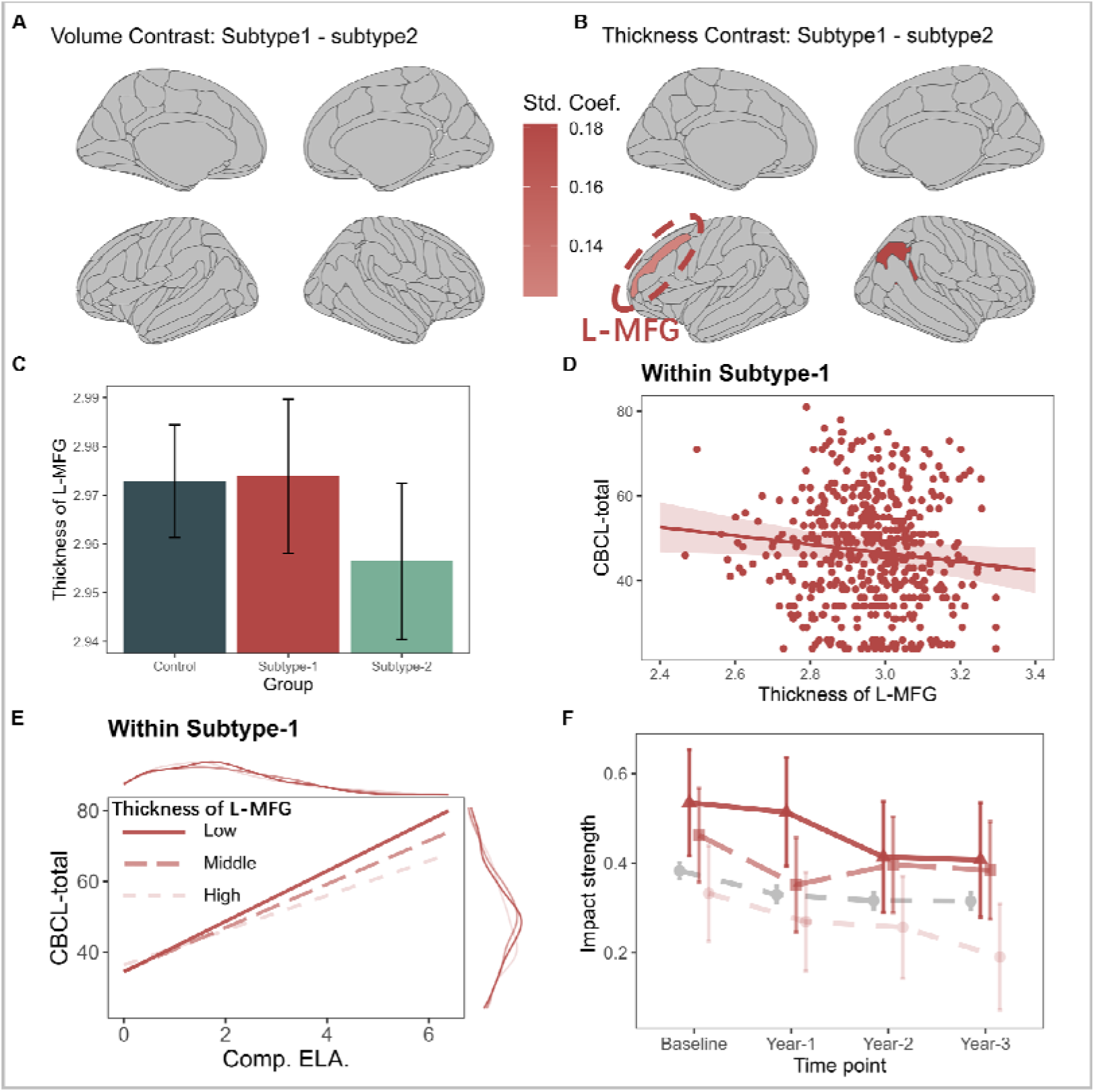
Modulation of cumulative ELAs’ impact on behavior problems by L-MFG thickness in poor children with subtype-1. **A and B,** regional differences in volume (A) and thickness (B) among poor children with heterogeneous neuroimaging phenotypes. **C,** differences in L-MFG thickness between groups (represented by different colors), with error bars indicating the 95% CIs. **D**, scatter plot of L-MFG thickness and CBCL-total score, with the best-fitting line and 95% CI (shaded area). **E**, differences in cumulative ELAs’ impact among groups, stratifying poor children with subtype-1 on L-MFG thickness (transparent and sparse lines represent children with lower thickness) into three groups. Variable distributions are visualized by density plots on the edges. **F.** trends of the impact strength of baseline-measured cumulative ELAs on CBCL-total score over time between groups.

### Relating activation maps of subtype-1/2 to neurotransmitter receptor/transporter maps

We investigated the association between neurotransmitter substrates and the two resilience mechanisms identified in our study (i.e., activation maps of subtype-1 and subtype-2, contrasted with control group). We used whole-brain distribution maps of neurotransmitter receptors and transporters derived from 36 studies, covering 19 receptors and transporters from approximately 1,200 healthy individuals (Hansen et al., 2022; https://github.com/canlab/Neuroimaging_Pattern_Masks/tree/master/Atlases_and_parcellations/2022_Hansen_PET_tracer_maps). These distribution maps were parceled into the Destrieux atlas (excluding subcortical regions; Destrieux et al., 2010). Pearson’s correlation coefficients between these maps and the activation maps of the two subtypes were calculated. The significance of these coefficients was determined through 10000 permutation tests, and spatial autocorrelation was controlled using the spin test (utilizing the gen_spinsamples function from the netneurotools package, https://github.com/netneurolab/netneurotools; Alexander-Bloch et al., 2018).

### Relating activation maps of subtype-1/2 to gene expression maps and enrichment analysis

The Allen Human Brain Atlas (AHBA; https://human.brain-map.org/) dataset includes high-resolution spatial genomic transcriptional data from the cortex of six postmortem brains (male/female = 4/2, mean age = 45 years). We preprocessed the AHBA data using the get_expression_data function from the abagen toolkit (default settings; https://github.com/rmarkello/abagen) and parceled the integrated data into the Destrieux atlas. Due to missing data in the right hemisphere for most subjects, we only utilized data from the left hemisphere. Two regions of interest (ROIs) with no data points were excluded, resulting in a matrix of 15,632 genes expressed across 72 regions.

We employed partial least squares (PLS) regression to predict the activation of 72 brain regions based on the 15,632*72 gene expression matrix, modeling subtype-1 and subtype-2 separately. The first PLS component (PLS1) was used for subsequent analysis. If the variance explained by PLS1 exceeded 95% of the variance explained by PLS1 in the null model (obtained via spin test), the co-expression of the 15,632 genes was considered significantly associated with the target activation map. In this study, only the PLS1 model for subtype-2 showed significant association.

For the significant PLS1 genes, we conducted enrichment analysis by categorizing them into one of seven major cell types in the central nervous system (excitatory neurons, inhibitory neurons, endothelial cells, astrocytes, microglia, oligodendrocytes, and oligodendroglial precursor cells), based on results from a meta-analysis (Seidlitz et al., 2020). We calculated the median rank of each set of cell-typical genes. A cell type was considered significantly positively or negatively enriched if the median ranks of its typical genes were greater or less than 95% of the median ranks derived from the null model obtained via the spin test.

### Sensitivity analysis

We did several additional analyses to test the robustness of our results. First, we repeated the analysis with only the participants with complete observations (excluding all the participants with any missing value) to rule out the confounding effect of missing value imputation. Then, to comprehensively assess the cumulative effect of all types of ELAs, we included all 14 ELAs in calculating cumulative ELAs, rather than just the 8 ELAs showing significant amplification effects. Finally, to test the sensitivity of the results to the potential impact of family-level effect between children from the same family (Daníelsdóttir et al., 2024), for families with more than one child participating in the study (siblings), we randomly selected one sibling from every sibling pair.

## Results

The sample included 10112 children. Mean (SD) age was 119.04 (7.5) months at baseline. A total of 4893 participants (48.4%) were female; 1492 (14.8%) had an income below the poverty threshold and was therefore defined as poor children. The mean (SD) score of CBCL-total was 45.84 (11.19). The mean (SD) score of cumulative ELAs (of 14 kinds) was 3.1 (1.5). There were 7401 (73.2%) participants who passed fMRI QC. Details are presented in Table 1.

**Table 1.**
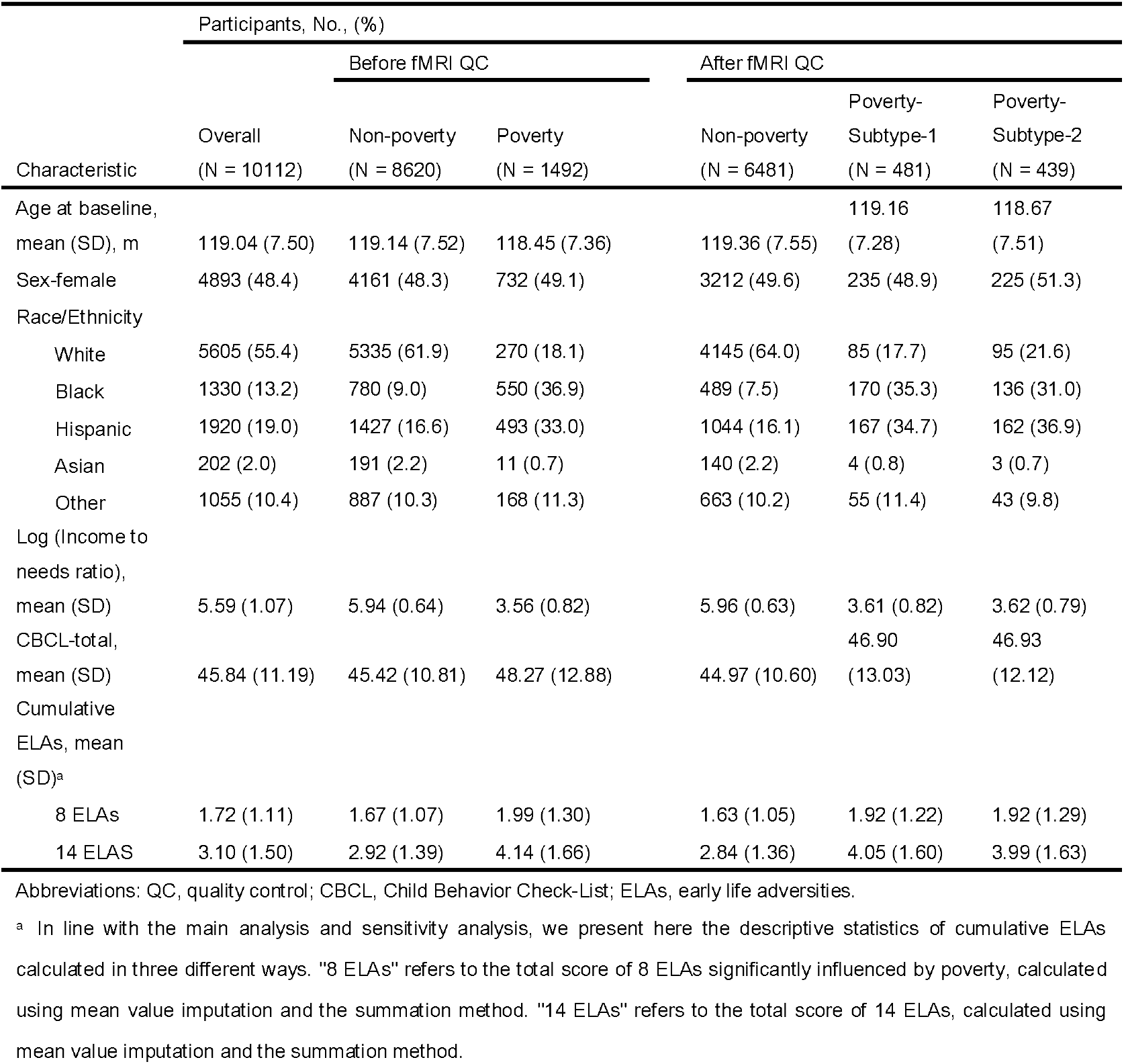
Characteristics of Study Participants.

### Poverty’s far-reaching and enduring impact on childhood resilience against various of behavior problems

We examined whether childhood poverty amplified the impact of ELAs on mental health. Our analysis revealed that childhood poverty increased the impact strength of 13 out of 14 ELAs on children’s behavior problems, with 8 ELAs showing significant increase effects. These significantly impactful ELAs include experience of surgery, prenatal exposure, experience of trauma, parent has substance abuse problem, sleep problem, experience of being bullied, family conflict, and ASR-total. For detailed statistical findings, including standardized regression coefficients illustrating ELAs’ impact strength on behavior problems within both poverty and control (non-poverty) groups, as well as comparisons of these coefficients and their significance level, refer to Supplementary Material S5. Additionally, comparisons (ratios) of standardized regression coefficients between poor and non-poor children are presented in Figure 1A. These findings underscore that childhood poverty exacerbates the impact of most ELAs on behavior problems, indicating disrupted resilience among poor children facing multiple kinds of risk factors.

Next, to investigate whether childhood poverty undermines resilience against the cumulative effect of multiple ELAs, we assessed the combined impact of 8 ELAs influenced significantly by poverty. Our analysis uncovered that compared to their non-poor counterparts, the impact strength of cumulative ELAs on behavior problem was greater in poor children (β_poor_ (SE) = 0.485 (0.018), β_non-poor_ (SE) = 0.397 (0.009); Δβ (SE) = 0.088 (0.02), *p* < .0001; Figure 1B). These findings suggest that in the context of cumulative ELAs, poor children demonstrate disrupted resilience.

Then, we further investigated whether the disruption of resilience in poor children persisted into adolescence. Analyzing the impact of cumulative ELAs across four waves, we found that childhood poverty continued to amplify this impact. Pooled longitudinal data showed significantly higher impact strength of baseline-measured cumulative ELAs on behavior problems in poor children compared to their non-poor counterparts over four waves (β_poor_ (SE) = 0.409 (0.016), β_non-poor_ (SE) = 0.336 (0.008); Δβ (SE) = 0.073 (0.018), *p* < .0001; Figure 1C). This impact remained significantly higher even after 3 years (β_poor_ (SE) = 0.375 (0.02), β_non-poor_ (SE) = 0.315 (0.009); Δβ (SE) = 0.073 (0.02), *p* = .0082; Figure 1C). Our analysis also revealed that almost all dimensions of behavior problems (eight syndrome subscales of CBCL) were more susceptible to cumulative ELAs in poor children (Figure 1D). Detailed statistical findings across the four waves can be found in Supplementary Material S6. These results indicate that disrupted resilience against cumulative ELAs in poor children persists into adolescence and also shape a broad range of behavior problems.

### Neuroimaging phenotypes distinguish intact and disrupted resilience among poor children

The HYDRA algorithm identified two heterogeneous subtypes of neural representations of self-regulation among poor children (see Supplementary Material S4 for details). During the stop-signal task, compared with the non-poor counterparts, poor children with subtype-1 exhibited increased activation across the entire brain (Figure 2A), while poor children with subtype-2 demonstrated decreased activation across the entire brain (Figure 2B). The greatest increases in activation between poor children with subtype-1 and their non-poor counterparts were observed in brain regions such as R-superior-parietal-lobule, R-middle-frontal-gyrus, R-lingual-gyrus, R-transverse-parietal-sulci, and L- superior-parietal-lobule. The greatest decreases in activation between poor children with subtype-2 and their non-poor counterparts were observed in brain regions such as R-superior-temporal-sulcus, L-precentral-gyrus, R-precentral-gyrus, R-inferior-frontal-sulcus, R- intraparietal-sulcus-and-transverse-parietal-sulci. Detailed statistical findings regarding these comparisons can be found in Supplementary Material S7.

In terms of resilience against cumulative ELAs, poor children with subtype-1 showed significantly higher impact strength of baseline-measured cumulative ELAs on behavior problems compared to their non-poor counterparts (β_poor-subtype-1_ (SE) = 0.533 (0.032), β_non-poor_ (SE) = 0.384 (0.01); Δβ (SE) = 0.149 (0.033), *p* < .0001; Figure 2C), whereas this difference was not significant in poor children with subtype-2 (β_poor-subtype-2_ (SE) = 0.433 (0.031); Δβ (SE) = 0.049 (0.033), *p* = .135; Figure 2C). Even after 2 years, the impact strength remained significantly higher for poor children with subtype-1 (β_poor-subtype-1_ (SE) = 0.412 (0.034), β_non-poor_ (SE) = 0.302 (0.011); Δβ (SE) = 0.11 (0.035), *p* = .0126; Figure 2D), but not for poor children with subtype-2 (β_poor-subtype-2_ (SE) = 0.37 (0.033); Δβ (SE) = 0.068 (0.035), *p* = .1194; Figure 2D). Detailed statistical findings are presented in Table 2 (baseline) and in Supplementary Material S8 (longitudinal analysis over 4 waves). These results suggest that neuroimaging phenotypes related to self-regulation are associated with intact or disrupted resilience among poor children against cumulative ELAs.

**Table 2.** Impact strength of cumulative ELAs on CBCL-total.

| CBCL-total ~<br>cumulative ELA<sup>a</sup> | $\beta$ (95% CI) | | | | |
| --- | --- | --- | --- | --- | --- |
|  | Before fMRI QC |  | After fMRI QC |  |  |
|  | Non-poverty | Poverty | Non-poverty | Poverty-Subtype-1 | Poverty-Subtype-2 |
| CBCL-total,<br>Baseline<sup>b</sup> |  |  |  |  |  |
| 8 ELAs | .397 (.379, .414) | .485 (.45, .520) | .384 (.364, .404) | .533 (.471, .595) | .433 (.372, .494) |
| 14 ELAs | .312 (.298, .326) | .381 (.353, .408) | .299 (.283, .316) | .413 (.365, .461) | .333 (.284, .382) |
| CBCL-total, Year-<br>1<sup>b</sup> |  |  |  |  |  |
| 8 ELAs | .330 (.312, .348) | .408 (.371, .444) | .322 (.301, .343) | .431 (.366, .495) | .397 (.332, .461) |
| 14 ELAs | .270 (.256, .285) | .322 (.293, .351) | .262 (.245, .279) | .334 (.285, .384) | .308 (.257, .360) |
| CBCL-total, Year-<br>2<sup>b</sup> |  |  |  |  |  |
| 8 ELAs | .316 (.298, .334) | .392 (.355, .429) | .302 (.281, .323) | .412 (.346, .478) | .370 (.306, .435) |
| 14 ELAs | .252 (.237, .267) | .292 (.263, .322) | .240 (.223, .257) | .295 (.245, .346) | .283 (.232, .335) |
| CBCL-total, Year-<br>3<sup>b</sup> |  |  |  |  |  |
| 8 ELAs | .315 (.297, .334) | .375 (.337, .414) | .315 (.293, .336) | .382 (.314, .45) | .376 (.309, .444) |
| 14 ELAs | .247 (.232, .262) | .293 (.263, .323) | .243 (.226, .261) | .297 (.245, .349) | .296 (.243, .350) |

Abbreviations: QC, quality control; CBCL, Child Behavior Check-List; ELAs, early life adversities.
<sup>a</sup>In line with the main analysis and sensitivity analysis, we present here the descriptive statistics of cumulative ELAs calculated in three different ways. "8 ELAs" refers to the total score of 8 ELAs significantly influenced by poverty, calculated using mean value imputation and the summation method. "14 ELAs" refers to the total score of 14 ELAs, calculated using mean value imputation and the summation method.
<sup>b</sup>The results at baseline were modeled based on the baseline data. The results at Year-1, Year-2, and Year-3 were modeled based on the data across the four years, with subject ID included as an additional random effect.

### Characteristics of heterogeneous neuroimaging phenotypes of poor children

#### No significant difference in superficial characteristics

To reveal the factors associated with the differentiation of neuroimaging phenotypes among poor children, we examined the characteristic differences in demographic, ELAs, mental health, and cognitive function between the poor children with heterogeneous neuroimaging phenotypes. No significant differences were observed in demographic characteristics (e.g., gender, race/ethnicity, family income), ELAs (as shown in Figure 1A), mental health (CBCL-total-score and subscales), or cognitive function (e.g., inhibitory control). Detailed statistical findings can be found in Supplementary Material S9. These results suggest that the differentiation of neuroimaging phenotypes among poor children may be driven by some more subtle factors.

#### Difference in brain morphometry

In terms of brain morphometry, we did not find any significant difference after multiple-comparison correction (*p* < .05, FDR correction). Therefore, for exploratory purposes, we report the results here without multiple comparison correction using a slightly stricter significance level (*p* < .01). Poor children with subtype-1 exhibited increased cortical thickness in brain regions including L-middle-frontal-gyrus (L-MFG; mean (SE) = 2.974 (0.008), Figure 3B) and R-angular-gyrus (R-AG; mean (SE) = 3.008 (0.009)), compared to poor children with subtype-2 (L-MFG, mean (SE) = 2.974 (0.008), Δmean = 0.01741, t = 2.78, *p* = .0054, Figure 3C; R-AG, mean (SE) = 2.979 (0.009), Δ = 0.029, t = 3.795, *p* = .0001). No significant difference was found between the two subtypes in the volumes of any regions (cortical or subcortical regions).

#### L-MFG’s resilience role exclusively within poor children with subtype-1

Given the widely established relationship between L-MFG and resilience (Brosch et al., 2022; Burt et al., 2016; Carballedo et al., 2012; Mary et al., 2020; Rodman et al., 2019; Salehinejad et al., 2017), we further explore the potential role of L-MFG in resilience. First, we discovered that higher L-MFG thickness was significantly associated with fewer behavior problems only within poor children with subtype-1 (β (SE) = -0.108 (0.053), *p* = .0411; Figure 3D), but not within poor children with subtype-2 (β (SE) = 0.013 (0.059), *p* = .8249) or non-poor children (β (SE) = 0.028 (0.015), *p* = .0752).

We then examined whether L-MFG thickness provided resilience against the cumulative effect of multiple ELAs. There was no significant interaction effect between cumulative ELAs and L-MFG thickness on CBCL-total score (β (SE) = -0.0369 (0.038), *p* = .3376) within poor children with subtype-1. However, when stratifying poor children with subtype-1 into three groups based on L-MFG thickness, the resilience role of L-MFG was revealed. Specifically, compared to children with the lowest L-MFG thickness, those with the highest L-MFG thickness demonstrated significantly lower impact strength of cumulative ELAs on behavior problems (β_high_ (SE) = 0.377 (0.053), β_low_ (SE) = 0.548 (0.058); Δβ (SE) = 0.171 (0.077), *p* = .0268; Figure 3E, solid line vs sparse dashed line). Even after 3 years, the impact strength remained significantly lower for individuals with the highest thickness (β_high-thick_ (SE) = 0.191 (0.06), β_low-thick_ (SE) = 0.408 (0.065); Δβ (SE) = 0.217 (0.088), *p* = .0498; Figure 3F). These findings highlight the resilient role of L-MFG against cumulative ELAs exclusively in poor children with subtype-1.

#### Neurochemical annotations and genomic signatures related to neuroimaging phenotypes

We further analyzed the molecular-level mechanisms underlying the heterogeneous neuroimaging phenotypes in poor children. We calculated the spatial similarities between gene expression and neurotransmitter maps and the fMRI patterns of the two neuroimaging phenotypes. Non-parametric two-sided permutation test controlling for spatial autocorrelation (spin test) was used to assess the significance of the similarities.

Among the 36 types of neurotransmitter receptors/transporters, the fMRI pattern of subtype-1 was significantly associated the neurotransmitter receptor/transporter distributions of 5HT (5-hydroxytryptamine) receptors 1a (5HT1a_1, *r* = .2, *p* = .01), and dopamine receptor D2 (D2_1, *r* = .18, *p* = .03; D2_3, *r* = .21, *p* = .01; see Fig. 4B and Supplementary Material S10 for details). There was no significant association between the fMRI pattern of subtype-1 and any type of neurotransmitter receptor/transporter distribution. These results suggested the potential link between neurochemical system and the disrupts resilience in poor children.

**Figure 4.**
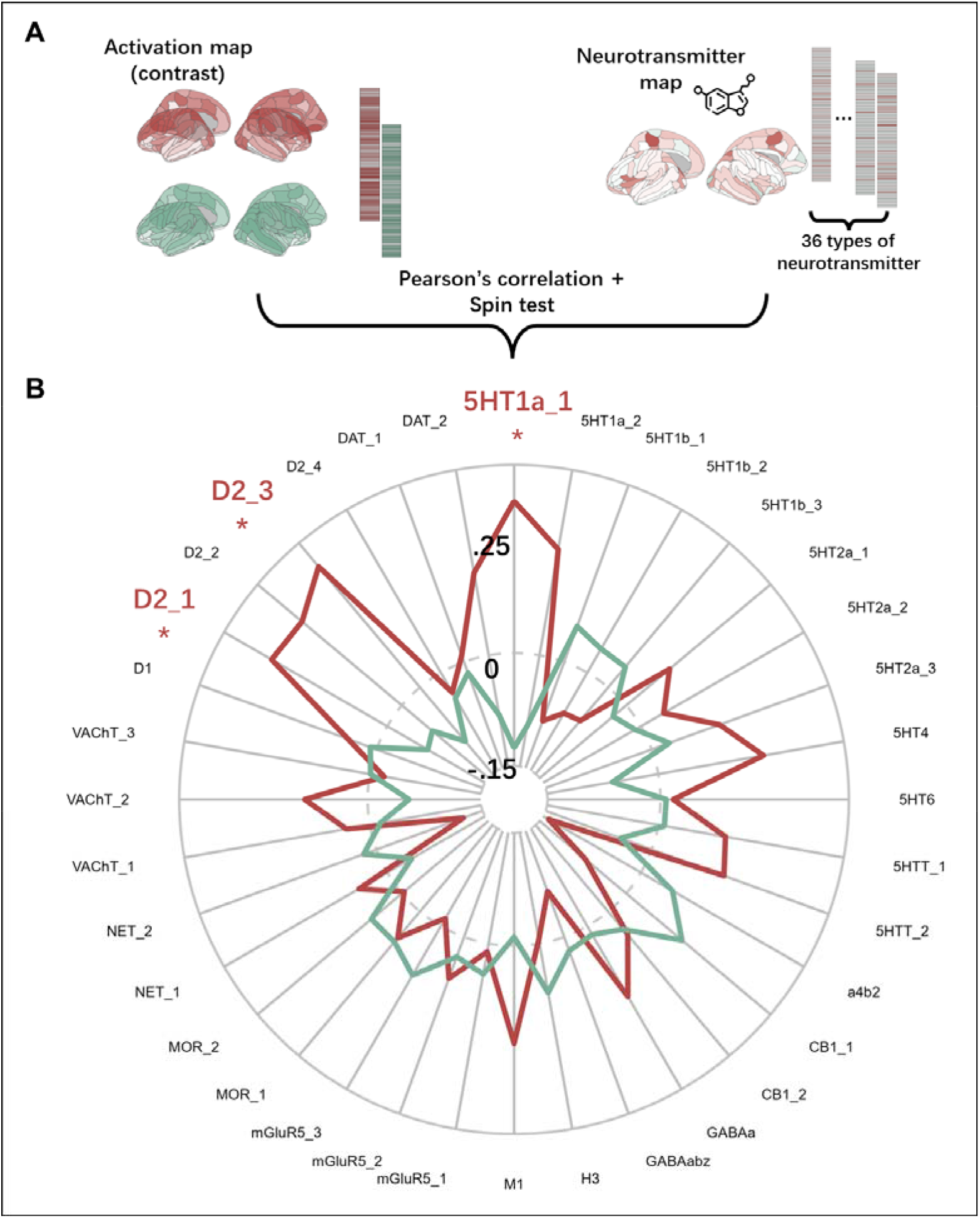
Spatial similarity between neurotransmitter receptor/transporter maps and activation maps of subtype-1/2. **A,** we parceled 36 different neurotransmitter receptor/transporter maps into 148 regions of interest (ROIs) using the Destrieux atlas. This resulted in 36 vectors representing the distribution of neurotransmitter receptors/transporters across the 148 ROIs. We then calculated the correlation coefficients between these vectors and the activation maps of subtype-1 and subtype-2 (contrasted with the control group), and assessed the significance using the spin test. **B,** radar plot illustrating the correlation coefficients between neurotransmitter receptor/transporter maps and activation maps of subtype-1 (shown in red) and subtype-2 (shown in green). Abbreviation: 5HT, 5-hydroxytryptamine. 5HTT, 5-hydroxytryptamine transporter. a4b2, alpha-4 beta-2 nicotinic receptor. CB1, cannabinoid receptor. D1, D2, dopamine receptors. DAT, dopamine transporter. GABA, γ-Aminobutyric acid receptor. H3, histamine receptor. M1, Muscarinic acetylcholine receptor. mGluR, metabotropic glutamate receptor. MOR, Mu opioid receptors. NET, norepinephrine transporter. VAChT, vesicular acetylcholine transporter.

PLS regression was used to characterize the relationship between gene expression (15632 genes) and fMRI patterns of the two neuroimaging phenotypes (PLS models were built separately for subtype-1 and subtype-2). The first PLS component (PLS1) explained a significant amount of covariance in subtype-2 (38.6% variance explained, *p* = .024, Fig. 5B), but not in subtype-1 (30.7% variance explained, *p* = .233, therefore no further analysis was performed). In subtype-2, the PLS1 gene expression weights were positively correlated with fMRI patterns. This means that genes with strongly negative loading on PLS1 (at the bottom of the ranked PLS1 list, like NAT8L gene in Fig. 5C), were overexpressed in regions with strongly negative activations. Then, gene sets were assigned to one of the seven cell types in the central nervous system (Seidlitz et al., 2020), and the median rank of genes in the PLS1 loadings within each cell types was calculated as an indicator of cell type enrichment. PLS1 exhibited significantly positive or negative enrichment across all seven cell types (*p* < .05, Fig. 5E). Significant positive enrichments were observed in astrocytes (median rank =3745 / 15632, *p* < .0001), oligodendrocytes (median rank = 4014 / 15632, *p* < .0001), microglia (median rank = 5565 / 15632, *p* < .0001), oligodendroglial precursor cells (OPC; median rank = 6480 / 15632, *p* < .0001). While the significant negative enrichments were observed in excitatory neurons (median rank = 10312 / 15632, *p* < .0001), inhibitory neurons (median rank = 9195.5 / 15632, *p* < .0001), endothelial cells (median rank = 8359.5 / 15632, *p* = .0189). These results suggested the potential link between gene expression profiles and the intact resilience in poor children.

**Figure 5.**
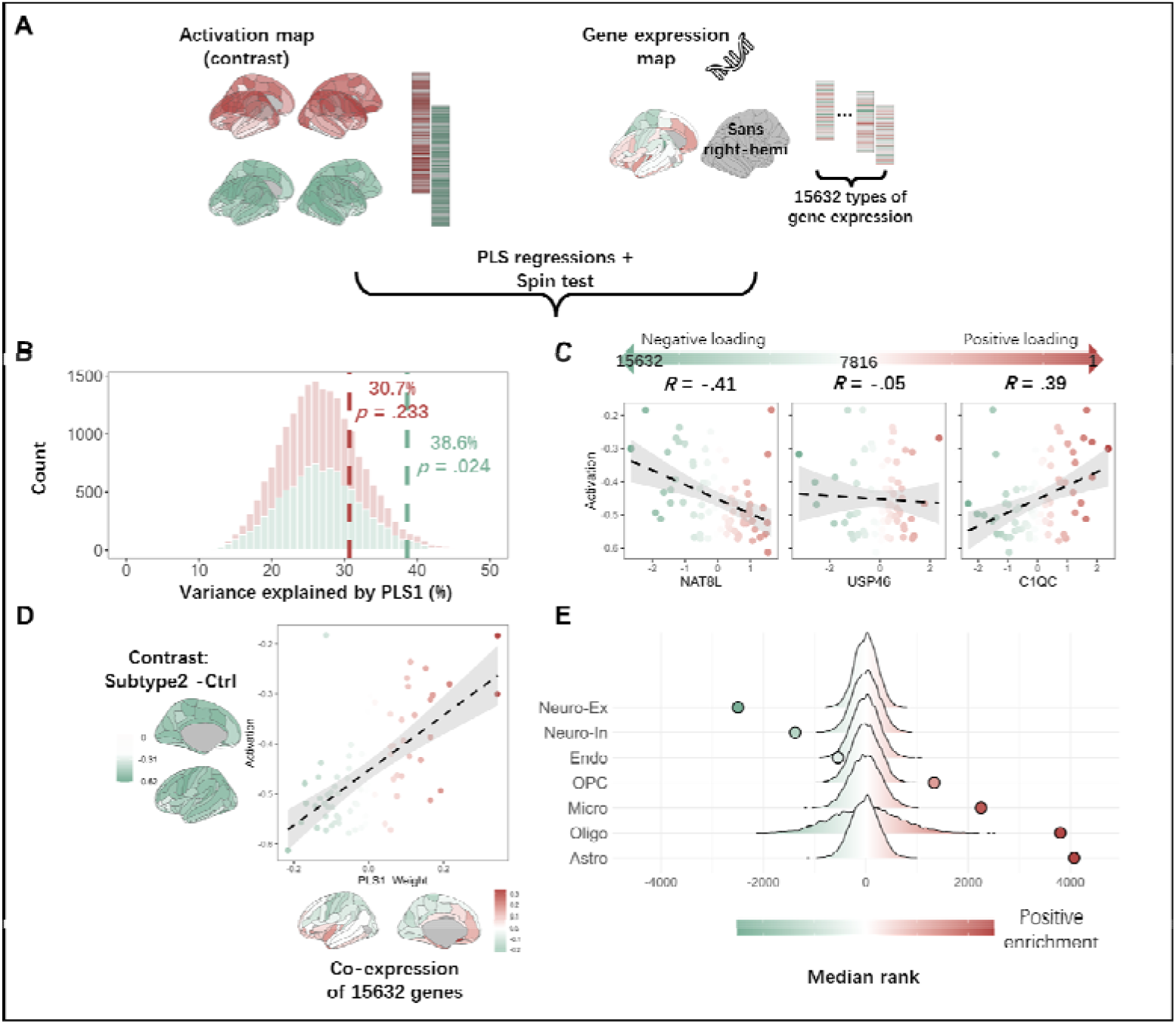
Spatial similarity between gene expression maps and activation maps of subtype-1/2. **A,** 15632 different gene maps were parceled into 72 regions of interest (ROIs) using the Destrieux atlas (left hemisphere only, excluding two ROIs with no data point), resulting in 15632 vectors representing gene expression distribution across the 72 ROIs. We performed PLS regression between the {15632 * 72} matrix of gene expression and the activation maps of subtype-1 and subtype-2 (contrasted with the control group) separately. The first PLS component (PLS1) was used for further analysis. **B,** the null distribution of the explained variance by PLS1 was derived from a spin test (10000 permutations, corrected for spatial autocorrelation) with the corresponding true value. Subtype-1 and subtype-2 are colored in red and green, respectively. **C,** the relationship between the activation map of subtype-2 and exemplary genes is shown. C1QC, a positively weighted gene near the top of the ranked PLS1 weights list; USP46, a near-zero weighted gene in the middle of the list; NAT8L, a negatively weighted gene near the bottom. Negatively weighted genes were more strongly expressed in regions with more deactivation (i.e., poor children with subtype-2 >> control group), whereas positively weighted genes were less strongly expressed in these regions. **D,** PLS1 was positively correlated with the activation map of subtype-2; thus, low PLS1 scores were correlated with more deactivation. **E,** cell type enrichment of the weighted, ranked gene list from PLS analysis of covariation between activation map of subtype-2 and gene expression is plotted. Median ranks of genes in each cell type are plotted in dot with their null distribution derived from the spin test. Negatively weighted genes were significantly enriched for genes expressed by excitatory neurons (Neuro-Ex), inhibitory neurons (Neuro-In), and endothelial cells (Endo). Positively weighted genes were significantly enriched for genes expressed by astrocytes (Astro), oligodendrocytes (Oligo), microglia (Micro), and oligodendroglial precursor cells (OPC).

### Sensitivity analysis

In sensitivity analyses, the difference in resilience against cumulative ELAs (impact strength of cumulative ELA on behavior problems) between groups (poverty vs non-poverty; non-poverty vs subtype-1 vs subtype-2; high-L-MFG vs low-L-MFG) remained the same when we included all 14 ELAs in calculating cumulative ELAs (rather than just the 8 ELAs showing significant amplification effects; see Supplementary Material S11), when we ruled out the confounding effect of missing value imputation (see Supplementary Material S12), and when we retained only one child for each family (see Supplementary Material S13).

## Discussion

In this study utilizing a nationwide population-based cohort of children, we found that various Early Life Adversities (ELAs) and their cumulative effects on behavioral problems are moderated by childhood poverty. Specifically, children from poor family exhibit a greater impact and cumulative effect of ELAs on behavioral problems compared to those from non-poor families, and this magnified effect persists into adolescence. That is, childhood poverty disrupts resilience. Furthermore, we further elucidated that this resilience disruption may be determined by the neural representation of self-regulation. Specifically, poor children with a certain phenotype of neural activation pattern of self-regulation show no difference in resilience compared to non-poor children, while those with another phenotype demonstrate disrupted resilience.

Consistent with numerous research findings, our study results suggest that reducing ELAs exposure remains a priority for promoting children’s mental health (Daníelsdóttir et al., 2024). Nevertheless, poor children are indeed more vulnerable/sensitive to the adverse impact of ELAs, resulting in disrupted resilience. Childhood poverty could disrupt resilience through allostatic overload, which refers to the wear and tear of the brain and body by excessive exposure to stressors (De France et al., 2022; B. S. McEwen et al., 2015). Specifically, the low socioeconomic status represented by poverty is accompanied by chronic and toxic stress, increasing children’s allostatic overload and leaving them with inadequate resources to cope with ELAs (C. A. McEwen & McEwen, 2017). Meanwhile, allostatic overload is further exacerbated by the high exposure to ELAs caused by childhood poverty, which encroaches the indebted resources of poor children, further compromising their resilience (Evans, 2004; C. A. McEwen & McEwen, 2017; Rakesh et al., 2023). Finally, poor children are less likely to minimize allostatic load with the help of a supportive environment (C. A. McEwen & McEwen, 2017; Rutter, 2012; Ungar & Theron, 2020). Therefore, when facing ELAs, poor children have more difficulty maintaining and restoring homeostasis, resulting in disrupted resilience.

However, it is still hopeful to nurture resilience amidst childhood poverty. Our findings indicate that heterogeneous neuroimaging phenotypes (related to self-regulation) are discriminative biomarkers of intact or disrupted resilience among poor children. It has become increasingly popular to subtype mental illness through neuroimaging phenotypes, providing significant insight for precision medicine and prediction of disease development (Brucar et al., 2023; Jiang et al., 2023; Zhang et al., 2023). Although poverty is not classified as a mental illness, it stands out as one of the most potent risk factors for many mental illnesses (Udalova et al., 2022) and plays a significant role in amplifying the effects of other risk factors (C. A. McEwen & McEwen, 2017). Therefore, subtyping childhood poverty and probing the potential biomarkers of intact resilience may offer crucial insights into the field of mental illness. Importantly, existing research has revealed the heterogeneity of the impact of childhood poverty, indicating that among poor children/adolescents, individuals with a higher level of self-regulation have better resilience against ELAs (Brody et al., 2013, 2020; De France et al., 2022; Ehrlich et al., 2024).

In this study, we found that poor children with better resilience exhibited lower whole-brain activation during the self-regulation task. This association can be explained by the neural efficiency hypothesis (Deary et al., 2010; L. Li & Smith, 2021; Neubauer & Fink, 2009), that efficient neural functioning in an EASY self-regulation task determines the successful self-regulation/adaption to more DIFFICULT ELAs in real-world environment. Conversely, this success would be relatively more difficult for individuals who require additional neural resources in EASY self-regulation task. Recent studies have revealed inefficient recruitment of the cognitive brain regions during self-regulation task in patients with trauma-induced mental illness (greater N2 amplitude and greater left inferior parietal activation measured by EEG and fMRI; Korgaonkar et al., 2021) and major depressive disorder (van Tol et al., 2011), in high-anxious individuals (greater dorsolateral prefrontal activation measure by fMRI; Basten et al., 2011), in adopted children at high genetic risk of depression (greater whole-brain activation measure by fMRI, using ABCD dataset; Petrican et al., 2023), and in problematic smartphone users (greater frontoparietal regions activation measure by fMRI; Choi et al., 2021). A recent meta-analysis has found that adversity exposure was associated with less efficient neural activity in prefrontal cortex across four neurocognitive domains (Hosseini-Kamkar et al., 2023). Importantly, after trauma exposure, better resilience was found to be associated with more efficient neural activity in key brain regions during sustained attention (Montalto et al., 2023). Our study supplements this literature by presenting findings derived from a sizable population-based cohort, which also provide valuable insights into the resilience mechanism of poor children confronting ELAs, particularly through the lens of holistic multivariate activation patterns across the entire brain, as opposed to isolated regional activations in previous studies (Mendes et al., 2022).

While poverty biologically embeds disrupted resilience in poor children’s brain, opportunities for fostering intact resilience persist. Our study identifies the left middle frontal gyrus (L-MFG) as a supplementary resilience mechanism exclusively among poor children with disrupted resilience (subtype-1). This finding not only corroborates prior literature on the significance of L-MFG in resilience (Brosch et al., 2022; Burt et al., 2016; Carballedo et al., 2012; Mary et al., 2020; Rodman et al., 2019; Salehinejad et al., 2017), but also underscores its compensatory role in resilience mechanism. Specifically, the resilience mechanism of L-MFG may function as a compensator solely in disadvantaged individuals with disrupted resilience (poor children showing inefficient neural activity), rather than in those with intact resilience (poor children showing efficient neural activity). By considering disrupted resilience itself as a challenge, this conditional resilience mechanism remains consistent with the core concept of resilience which entails organismic adaptation to challenges (; Karatsoreos & McEwen, 2013; B. S. McEwen & Gianaros, 2010; Nederhof & Schmidt, 2012; Rutter, 2006; Snell-Rood & Snell-Rood, 2020). Poverty represent one of the most complex situations, interwoven with diverse challenges and ELAs for children (C. A. McEwen & McEwen, 2017; Tooley et al., 2021; Tribble & Kim, 2019). Consequently, fostering resilience mechanisms in poor children proves exceptionally difficult and complex. Nonetheless, childhood marks a critical period characterized by profound psychological and neural transformations (McClelland & Cameron, 2012; Pas et al., 2021; Raffaelli et al., 2005), offering ample opportunities for cultivating heterogeneous resilience mechanisms, even among disadvantaged ones. Hence, based on our study’s findings, we advocate for heightened research focus on resilience against poverty and other ELAs. Such endeavors should transcend superficial resilience mechanism discovery and delve deeper into uncovering heterogeneous resilience mechanisms latent within the disadvantaged populations.

The two resilience mechanisms revealed in our study, moreover, may not compete or influence each other directly. Instead, the mechanism most suited to an organism’s unique profile and environmental context is likely to be embodied. Utilizing publicly available neurotransmitter and gene expression maps, we explored the origins of these mechanisms. We found that the resilience mechanism characterized by worse neural efficiency and intact L-MFG (subtype-1) appears to be driven by serotonin (5HT) and dopamine receptors, known for their roles in stress recovery and resilience (Holz et al., 2020; Homberg & Jagiellowicz, 2022; Shinohara et al., 2018, 2018). Conversely, the resilience mechanism characterized by better neural efficiency (subtype-2) appears to be driven by brain gene co-expression. Enrichment analysis showed these genes are most enriched in astrocytes (B. Li et al., 2022; S. Li et al., 2022; Murphy-Royal et al., 2020) and excitatory neurons, consistent with existing stress and resilience research (Nakamura et al., 2022; Shinohara et al., 2018). In summary, using neurotransmitter and gene expression maps has deepened our understanding of resilience mechanisms. Future research should use PET and transcriptome analysis to further explore these mechanisms’ molecular origins and their interaction with environmental variation (Nakamura et al., 2022). These efforts will further elucidate the development of stress-related mental illnesses and guide precision medicine in psychiatry.

### Limitations

This study has several limitations. First, the findings related to the L-MFG in this study have not been corrected by multiple comparisons. Although the L-MFG is well recognized for its significant role in resilience (as mentioned above), these results should still be interpreted with caution and replicated in independent samples. Second, the assessment of children’s mental health relied on parent-reported data. While the Child Behavior Checklist (CBCL) has proven reliability and validity (Barch et al., 2021; Moore et al., 2020), it remains susceptible to reporting biases, including parent-reported family income (Udalova et al., 2022). The use of self-report measures is often necessary in large-scale studies, but further validation in clinical samples is essential to substantiate these findings. Lastly, given the findings related to “skin-deep resilience (i.e., for poor children, persistently maintaining high self-regulation may damage physical health; Brody et al., 2013, 2020; De France et al., 2022; Ehrlich et al., 2024), it is imperative to subsequently investigate the long-term consequences of various resilience manifestations (to adulthood), particularly concerning physical health.

## Data Availability

Data is from the Adolescent Brain and Cognitive Development Study. Information on how to access ABCD data through the NIMH Data Archive (NDA) is available on the ABCD study data sharing webpage: https://abcdstudy.org/scientists/data-sharing/

## S1: Descriptions of early life adversity (ELA) measures

### S1.1: Experience of surgery (categorical)

In this study, parents were asked to report their children’s surgical history (scrn_surgery), “Has your child ever had surgery?”, “1 = Yes; 0 = No”

### S1.2: Prenatal Exposure (continuous)

Children’s prenatal exposure was measured following the instruction of a previous work (Roffman et al., 2021), which encompass 6 exposures which were independently associated with significant but small increases in CBCL total score. These exposures included:

(1) Unplanned pregnancy (devhx_6_p)
(2) Maternal alcohol use early in pregnancy (devhx_8_alcohol)
(3) Maternal marijuana use early in pregnancy (devhx_8_marijuana)
(4) Maternal tobacco use early in pregnancy (devhx_8_tobacco)
(5) Pregnancy complications (positive response to any item from devhx_10a3_p to devhx_10m3_p)
(6) Birth complications (positive response to any item from devhx_14a3_p to devhx_14h3_p)

The incidences of these exposures were summed (0 ∼ 6) and rescaled to a range of 0 to 1.

### S1.3: Experience of trauma (categorical)

Parents in this study reported whether either parent had a history of alcohol problems (famhx_ss_parent_alc_p) or drug use problems (famhx_ss_parent_dg_p). If at least one parent had either an alcohol or a drug use problem, the child was recorded as having a positive exposure.

### S1.4: Parent has substance abuse problem (categorical)

Parent reported whether any one of the child’s parent(s) had alcohol problem (famhx_ss_parent_alc_p) or drug use problem (famhx_ss_parent_dg_p). If at least one of the parent(s) had either alcohol or drug use problem, the child was recorded as positive exposure.

### S1.5: Parental separation (categorical)

Parents reported their marital status using the question (demo_prnt_marital_v2): "Are you now married, widowed, divorced, separated, never married, or living with a partner?" The response options were coded as follows: 1 = Married, 2 = Widowed, 3 = Divorced, 4 = Separated, 5 = Never married, 6 = Living with a partner. If the response was neither 1 (Married) nor 6 (Living with a partner), the child was recorded as having a positive exposure to parental separation.

### S1.6: Area deprivation index (continuous)

Area deprivation index was provided by the ABCD group directly (reshist_addr1_adi_perc). Details can be found at https://wiki.abcdstudy.org/release-notes/non-imaging/linked-external-data.html#area-deprivation-index-adi

### S1.7: Neighbourhood insafety (continuous)

Neighbourhood insafety was assessed using the mean score of three questions (nsc_p_ss_mean_3_items) regarding the neighbourhood environment: "I feel safe walking in my neighborhood, day or night," "Violence is not a problem in my neighborhood," and "My neighborhood is safe from crime." Responses were rated on a scale from 1 ("Strongly Disagree") to 5 ("Strongly Agree"). The scores were then rescaled to a range of 0 to 1.

### S1.8: Sleep problem (continuous)

Parents reported their children’s sleep problems by responding to the question: "In the past two weeks, how often did your child have trouble falling asleep or staying asleep when he or she was tired and wanted to sleep?" Responses were rated on a scale from 0 ("Not at all") to 4 ("Nearly every day"). The scores were then rescaled to a range of 0 to 1.

### S1.9: School-bad environment (continuous)

The school environment was assessed using the School Environment Subscale of the School Risk and Protective Factors Survey (srpf_y_ss_ses). This subscale includes 6 questions, such as "In my school, students have lots of chances to help decide things like class activities and rules," with responses scored from 1 to 4. The sum score was then rescaled to a range of 0 to 1.

### S1.10: Experience of being bullied (categorical)

Parents reported their child’s experience of bullying through a single question from the survey (kbi_p_c_bully): "Does your child have any problems with bullying at school or in your neighborhood?" Responses were coded as 1 = Yes and 0 = No.

### S1.11: Family Conflict (categorical)

Family conflict was measured using the conflict subscale of the Family Environment Scale (fes_p_ss_fc), which includes 9 questions. For example, one question is "We fight a lot in our family," with responses coded as 1 = True and 0 = False.

### S1.12: ASR-total (continuous)

ASR-total (asr_scr_totprob_t) represents the total problems reported by the primary caregiver as a measure of parent self-reported psychopathological syndromes using the Adult Self Report (ASR). These scores were transformed to T-scores based on national probability samples and then rescaled to a range of 0 to 1.

### S1.13 Parent neglect (continuous)

Parent neglect was assessed through children’s self-report using the mean score (pmq_y_ss_mean) of the Parental Monitoring Subscale in The Parental Monitoring Survey. This subscale encompasses 5 questions, such as "How often do your parents/guardians know where you are?", with responses scored from 1 ("Never") to 5 ("Always or Almost Always"). The scores were then rescaled to a range of 0 to 1.

### S1.14 Parent coldness (continuous)

Parent neglect was assessed through children’s self-report using the mean score (crpbi_y_ss_parent) of the Acceptance Subscale in the Children’s Report of Parental Behavioral Inventory. This subscale encompasses 5 questions, such as "First caregiver (caregiver participating in study/completing protocol). Makes me feel better after talking over my worries with him/her", with responses scored from 1 ("Not like him/her") to 3 ("A lot like him/her"). The scores were then rescaled to a range of 0 to 1.

## S2: Descriptions of cognitive function measures

Children’s cognitive function was measured by the NIH Toolbox, encompassing seven tasks administered via iPad. We used norm-referenced T-scores of the seven tasks and their composite scores (crystallized cognition score, fluid cognition score, and total score) for all the analysis. Details regarding the measurement and scoring can be found at https://wiki.abcdstudy.org/release-notes/non-imaging/neurocognition.html#nih-toolbox-cognition

**Picture Vocabulary task** was used to assess children’s language vocabulary knowledge (nihtbx_picvocab_fc). **Flanker Inhibitory Control & Attention task** was used to assess children’s attention, cognitive control, executive function, and inhibition of automatic response 2(nihtbx_flanker_fc). **Picture Sequence Memory task** was used to assess children’s episodic memory and sequencing (nihtbx_picture_fc). **Dimensional Change Card Sort task** was used to assess children’s executive functiohttps://wiki.abcdstudy.org/release-notes/imaging/task-fmri-behavior.htmln (including set shifting, flexible thinking, concept formation; nihtbx_cardsort_fc). **Pattern Comparison Processing Speed task** was used to assess children’s information processing and processing speed (nihtbx_pattern_fc). **Oral Reading Recognition task** was used to assess children’s language, oral reading (decoding) skills, and academic achievement (nihtbx_reading_fc). **List Sorting Working Memory task** was used to assess children’s working memory and information processing (nihtbx_list_fc).

**Crystallized cognition score** (nihtbx_cryst_fc) was an integration of Picture Vocabulary task and Oral Reading Recognition task. **Fluid cognition** score was an integration (nihtbx_fluidcomp_fc) of Flanker Inhibitory Control & Attention task, Picture Sequence Memory task, Dimensional Change Card Sort task, Pattern Comparison Processing Speed task, and List Sorting Working Memory task. **Total score** (nihtbx_totalcomp_fc) was an integration of all seven tasks.

## S3: Preprocessing and quality control of neuroimaging data

The preprocessing of the raw neuroimaging data was conducted by the ABCD study group, which was arranged into tabulated data for the convenience of researchers performing secondary analysis. Details regarding MRI information (scanner information, scanning parameters) can be found at https://wiki.abcdstudy.org/release-notes/imaging/overview.html. Details regarding the preprocessing of neuroimaging data are detailed described by Hagler et al. (2019), or can be found at the website mentioned above. The current study mainly depended on the tabulated data of **brain activation** (contrasting between correct-stop trails and correct-go trials) **during stop signal task (SST)** (merging the cortical parcellation of <u>mri_y_tfmr_sst_csvcg_dst.csv</u> and subcortical parcellation of <u>mri_y_tfmr_sst_csvcg_aseg.csv</u>), the tabulated data of **thickness** (merging the cortical parcellation of <u>mri_y_tfmr_sst_csvcg_dst.csv</u> and subcortical parcellation of <u>mri_y_tfmr_sst_csvcg_aseg.csv</u>), the tabulated data of **volumes** (merging the cortical parcellation of <u>mri_y_tfmr_sst_csvcg_dst.csv</u> and subcortical parcellation of <u>mri_y_tfmr_sst_csvcg_aseg.csv</u>), all downloaded from ABCD 5.0 data release.

Quality control (QC) of neuroimaging data was in line with the recommendation of ABCD study group. Two variables in the table of recommended image inclusion (mri_y_qc_incl.csv) was used as the QC criteria in our study, i.e., “imgincl_sst_include = 1” was used as the QC criteria of SST, “imgincl_t1w_include = 1” was used as the QC criteria of thickness and volume. Details regarding the QC criteria can be found at https://wiki.abcdstudy.org/release-notes/imaging/quality-control.html#recommended-imaging-inclusion, and are detailed described by Hagler et al. (2019).

## S4: The Heterogeneity Through Discriminative Analysis (HYDRA)

HYDRA, a non-linear semi-supervised machine learning algorithm, is used for integrated binary classification and subpopulation clustering without needing a priori cluster specification or relying on similarity measures affected by non-specific factors like age and sex (https://github.com/evarol/HYDRA; Varol et al., 2017). The algorithm classifies individuals based on deviation indices between clinical and healthy reference groups, using a convex polytope formed by multiple linear hyperplanes. This approach models potential heterogeneity within clinical samples and extends linear classifiers into non-linear spaces. HYDRA was applied to regional activation measures (beta weight for SST all correct stop versus correct go contrast in subcortical and cortical regions), with poverty and non-poverty as diagnostic label, while controlling for the effect of age, birth gender, head motion (mean framewise displacement in mm), race/ethnicity (dummied), study site (dummied) as covariates. We assessed HYDRA models with 2-5 clusters while applying 10-fold cross-validation, the best model was determined based on the Adjusted Rand Index (ARI). Details regarding the algorithm can be found in the original work by Varol et al. (2017).

Compared to other models, the model with 2 clusters showed highest ARI of 0.50486, indicating that the neural representation of self-control in poor children can be best (with highest stability) characterized by 2 classes.

We then examined the statistical significance of the model with 2 clusters via permutation test. The null models in the permutation test are generated by randomly shuffling the poverty/non-poverty label. 100 null models were generated. We found that the ARI of real model was significantly higher than the ARIs of the null models (*p* < 0.05). These results suggested the 3-cluster solution as the optimal subtyping model.

## S5: Association between poverty × ELAs and behavior problem

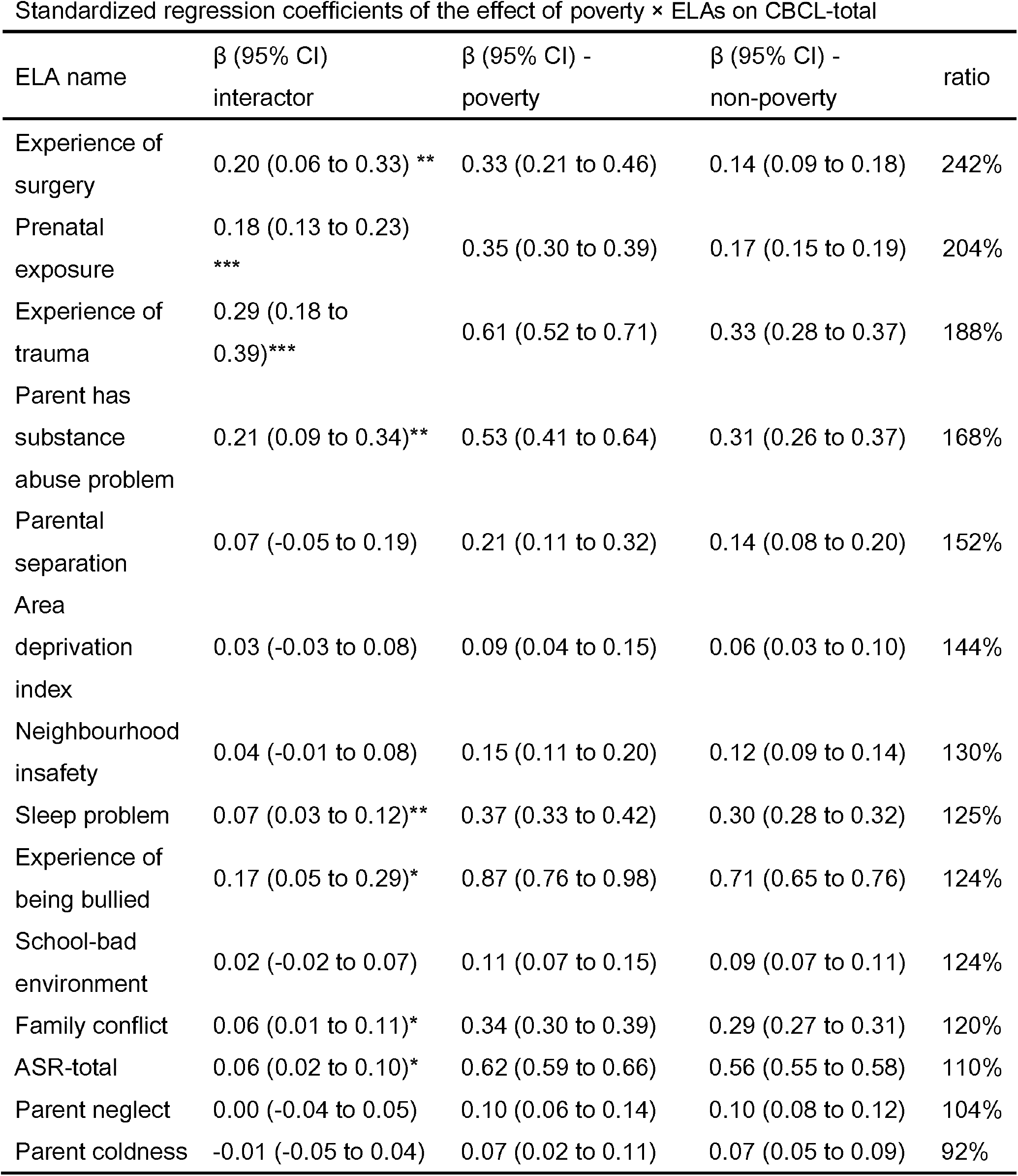
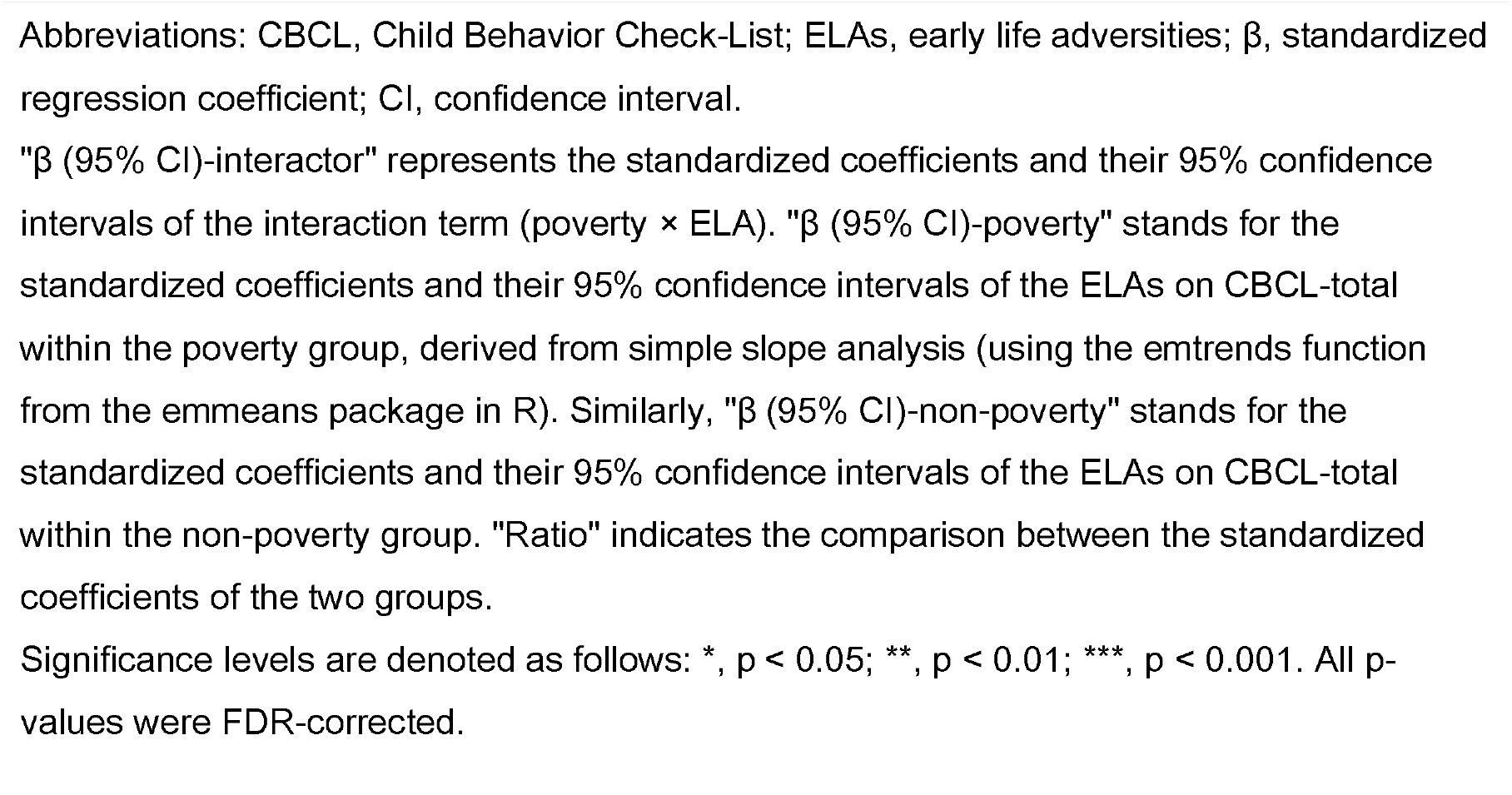

## S6: Longitudinal association between poverty × ELAs and various dimensions of behavior problem

| CBCL- | | $\Delta\beta$ (95% CI) | | | |
| --- | --- | --- | --- | --- | --- |
| total | Overall | Baseline | Year-1 | Year-2 | Year-3 |
|  | -0.15 (- | -0.19 (- | -0.16 (- | -0.13 (- | -0.13 (- |
| Aggres | 0.19 to - | 0.23 to - | 0.20 to - | 0.17 to - | 0.17 to - |
| sive | 0.12)* | 0.14)* | 0.12)* | 0.09)* | 0.08)* |
|  | -0.17 (- | -0.20 (- | -0.21 (- | -0.17 (- | -0.10 (- |
| RuleBr | 0.21 to - | 0.25 to - | 0.25 to - | 0.21 to - | 0.15 to - |
| eak | 0.14)* | 0.16)* | 0.17)* | 0.13)* | 0.06)* |
|  | -0.08 (- | -0.10 (- | -0.09 (- | -0.07 (- | -0.06 (- |
| Attentio | 0.12 to - | 0.14 to - | 0.13 to - | 0.11 to - | 0.11 to - |
| n | 0.04)* | 0.06)* | 0.05)* | 0.03)* | 0.02)* |
|  | -0.08 (- | -0.09 (- | -0.08 (- | -0.08 (- | -0.06 (- |
| Though | 0.11 to - | 0.13 to - | 0.12 to - | 0.12 to - | 0.11 to - |
| t | 0.04)* | 0.05)* | 0.03)* | 0.03)* | 0.02)* |
|  | -0.09 (- | -0.10 (- | -0.10 (- | -0.10 (- | -0.07 (- |
|  | 0.13 to - | 0.14 to - | 0.14 to - | 0.14 to - | 0.11 to - |
| Social | 0.06)* | 0.06)* | 0.06)* | 0.05)* | 0.02)* |
|  | -0.06 (- | -0.06 (- | -0.06 (- |  | -0.06 (- |
| Somati | 0.09 to - | 0.10 to - | 0.10 to - | -0.04 (- | 0.11 to - |
| c | 0.02)* | 0.01)* | 0.01)* | 0.09 to 0.00) | 0.02)* |
|  | -0.08 (- | -0.12 (- | -0.05 (- | -0.07 (- | -0.07 (- |
| WithDe | 0.11 to - | 0.16 to - | 0.10 to - | 0.11 to - | 0.12 to - |
| p | 0.04)* | 0.08)* | 0.01)* | 0.03)* | 0.02)* |
| AnxDE | -0.04 (- | -0.04 (- | -0.03 (- | -0.04 (- | -0.05 (- |
| p | 0.08 to - | 0.08 to 0.00) | 0.08 to 0.01) | 0.09 to 0.00) | 0.09 to - |
|  | 0.00)* |  |  |  | 0.00)* |
|  | -0.07 (- | -0.08 (- | -0.08 (- | -0.08 (- | -0.06 (- |
|  | 0.11 to - | 0.12 to - | 0.12 to - | 0.12 to - | 0.10 to - |
| **Total** | 0.04)* | 0.04)* | 0.04)* | 0.03)* | 0.02)* |

Statistics in the table were derived from simple slope analysis (using the emtrends function from the emmeans package in R). $\Delta\beta$ represents the comparison of $\beta$ between poor children and non-poor children. Comparisons were either made over four waves together (in the “Overall” column) or separately in each wave (from “Baseline” to “Year-3” columns).
Significance levels are denoted as follows: \*, $p < 0.05$ ; all p-values were FDR-corrected.

## S7: Group difference in brain activation during SST

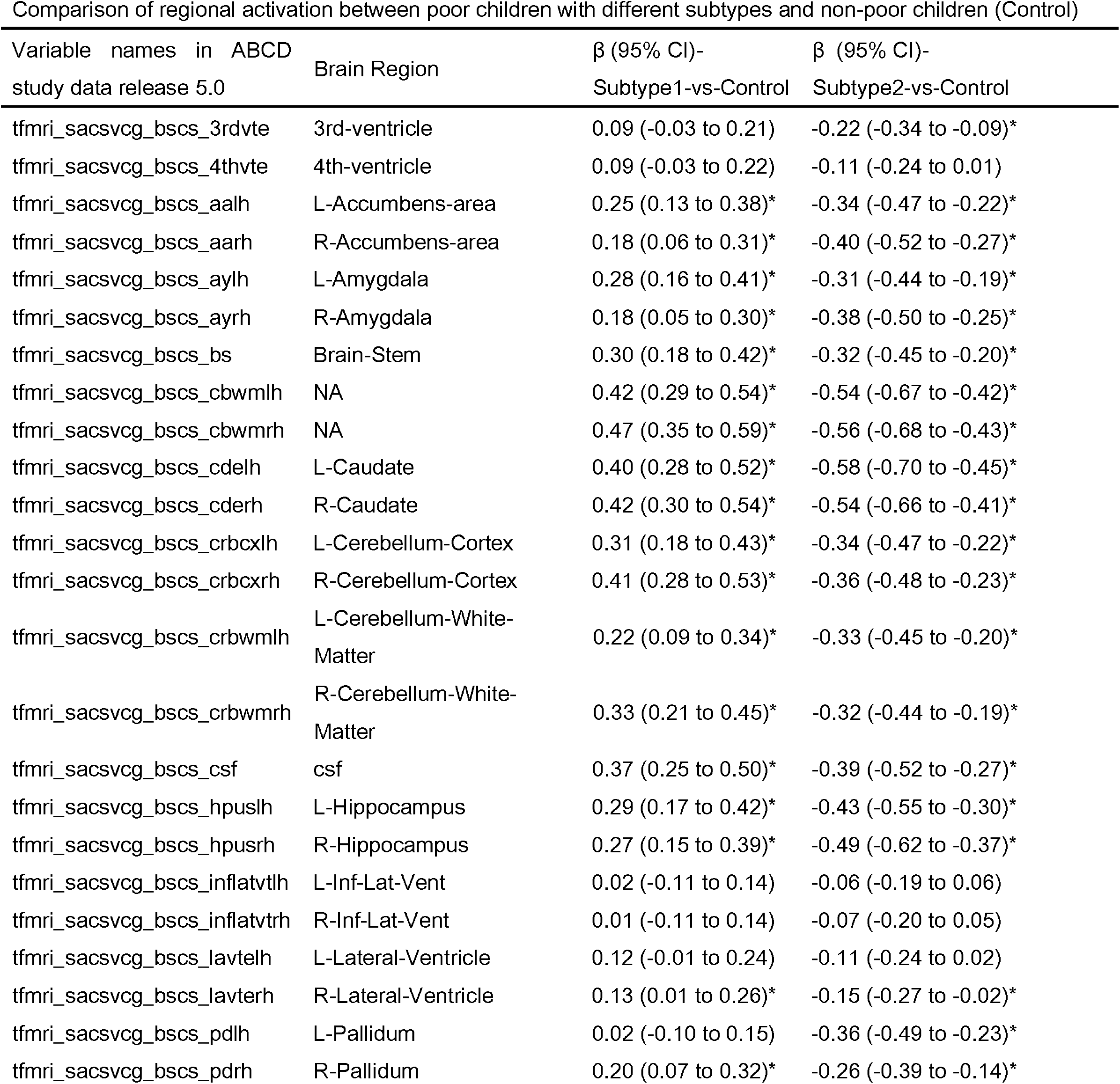
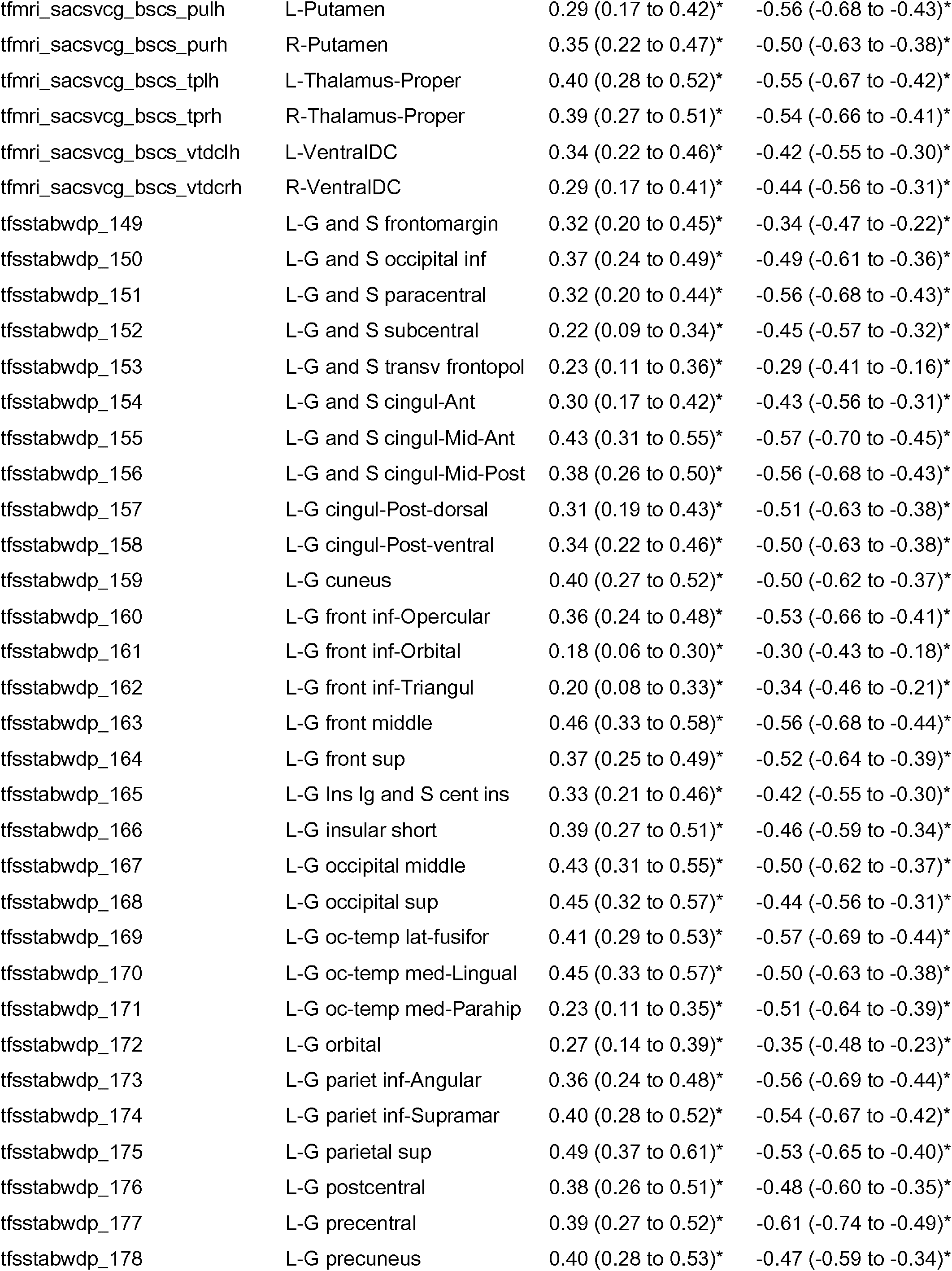
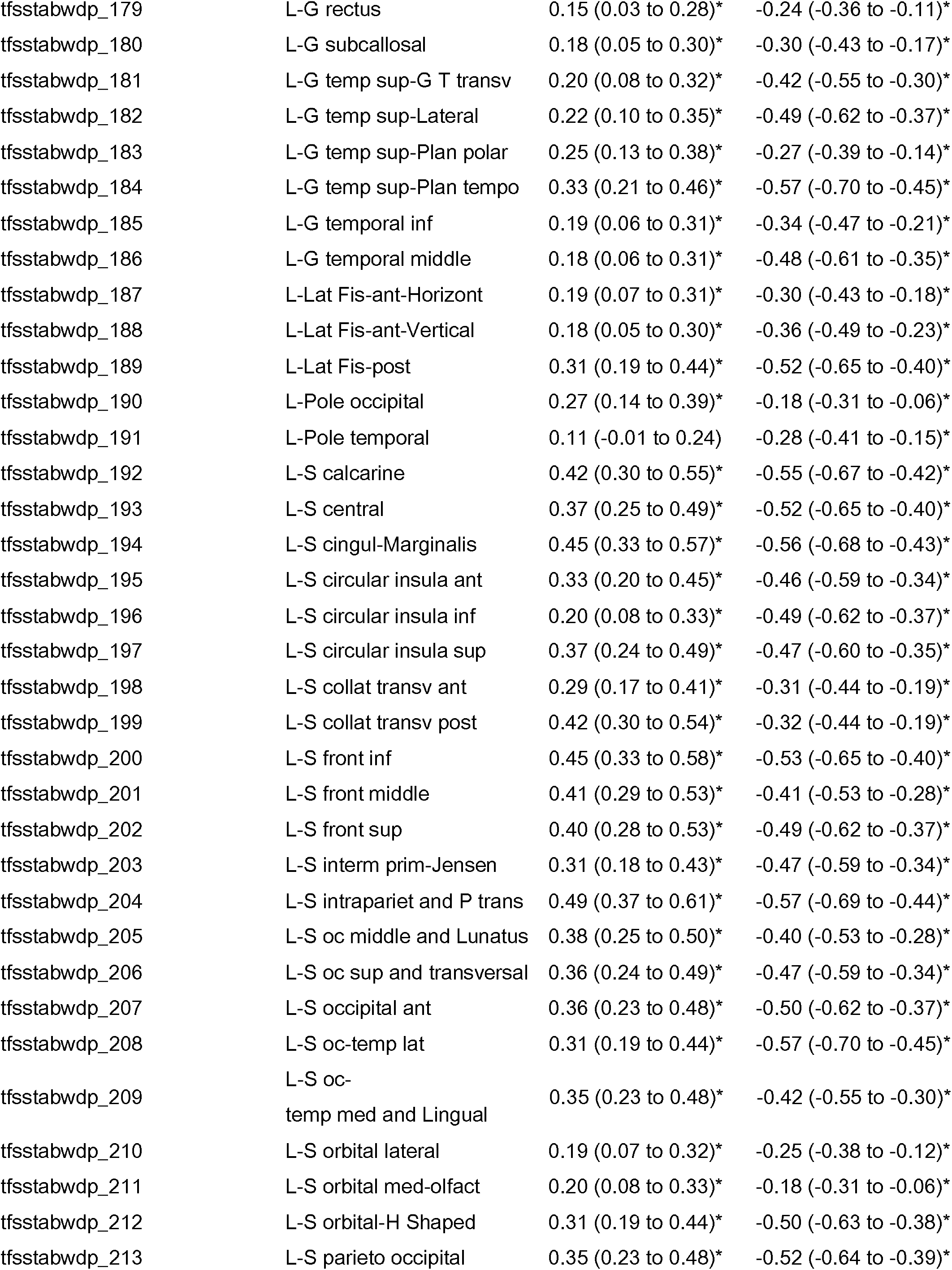
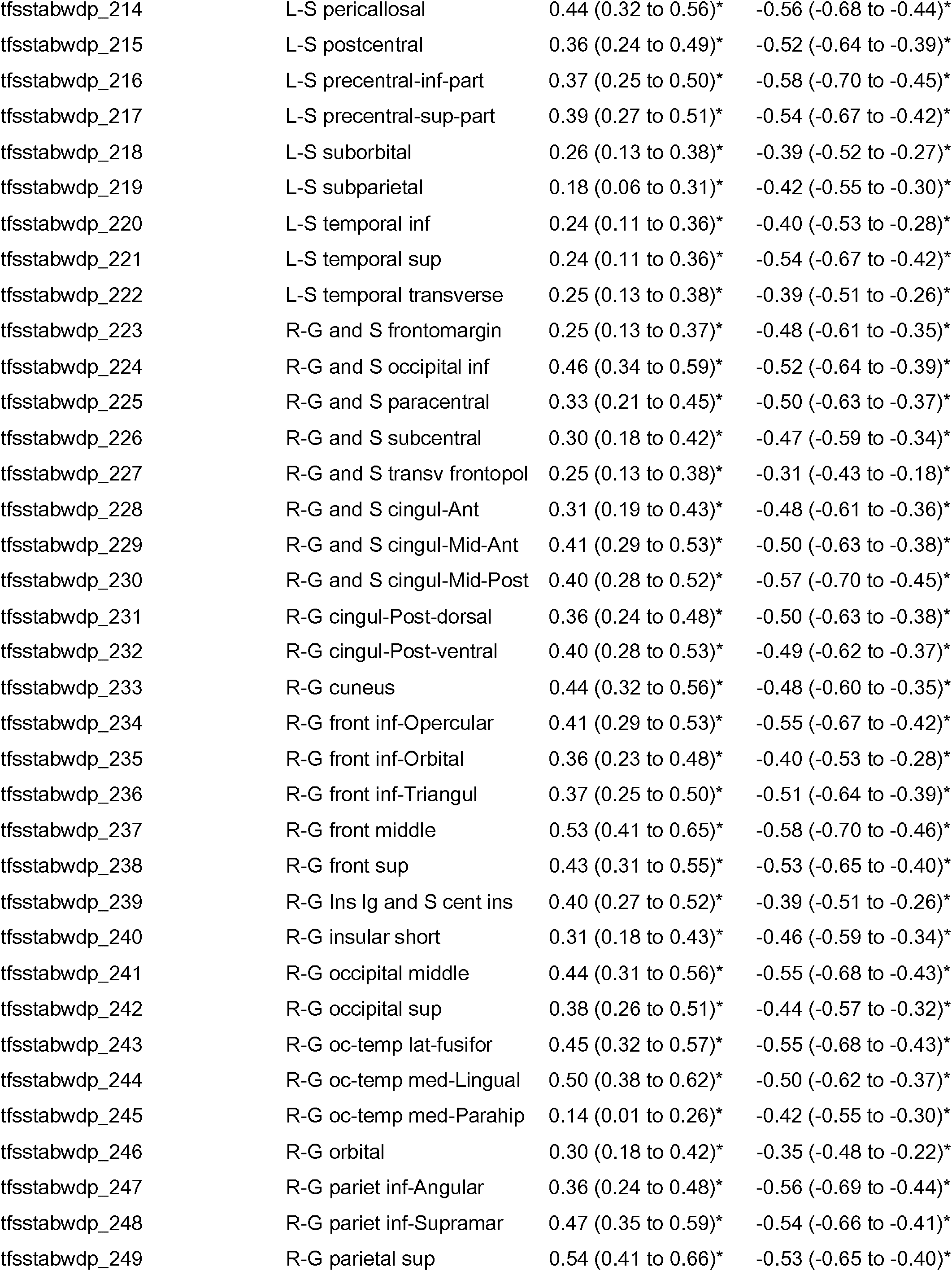
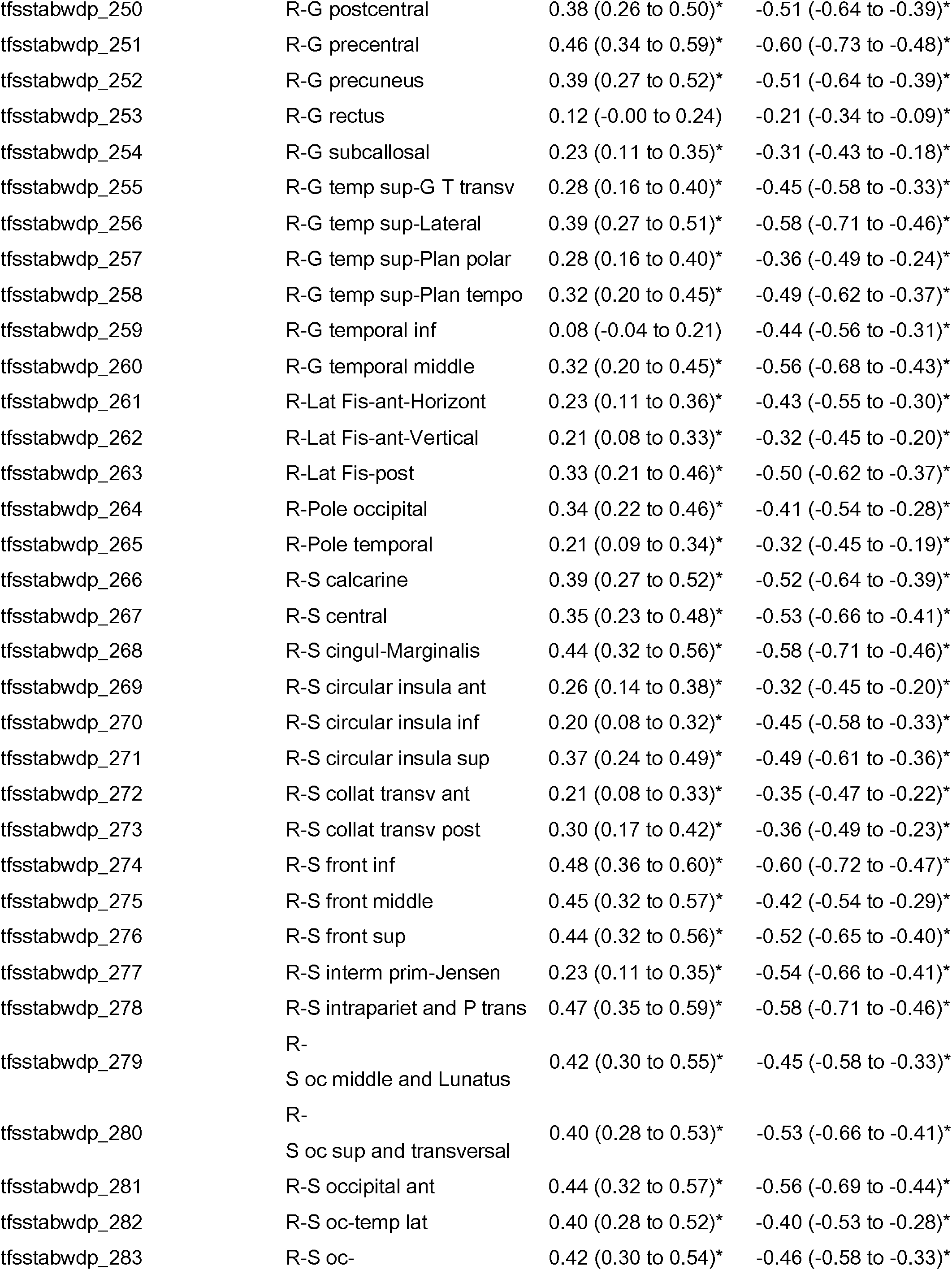
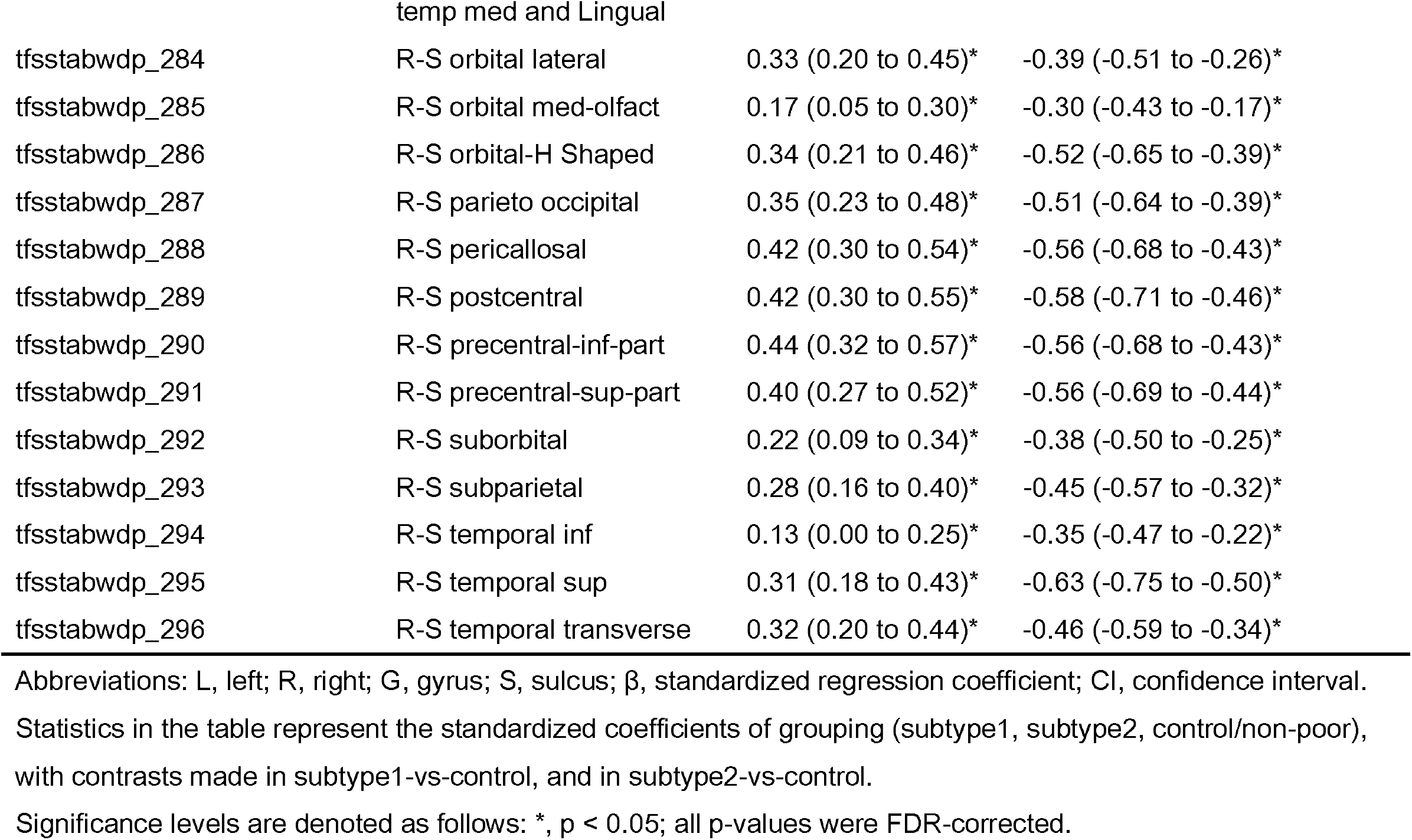

## S8: Longitudinal association between subtype × ELAs and various dimensions of behavior problem

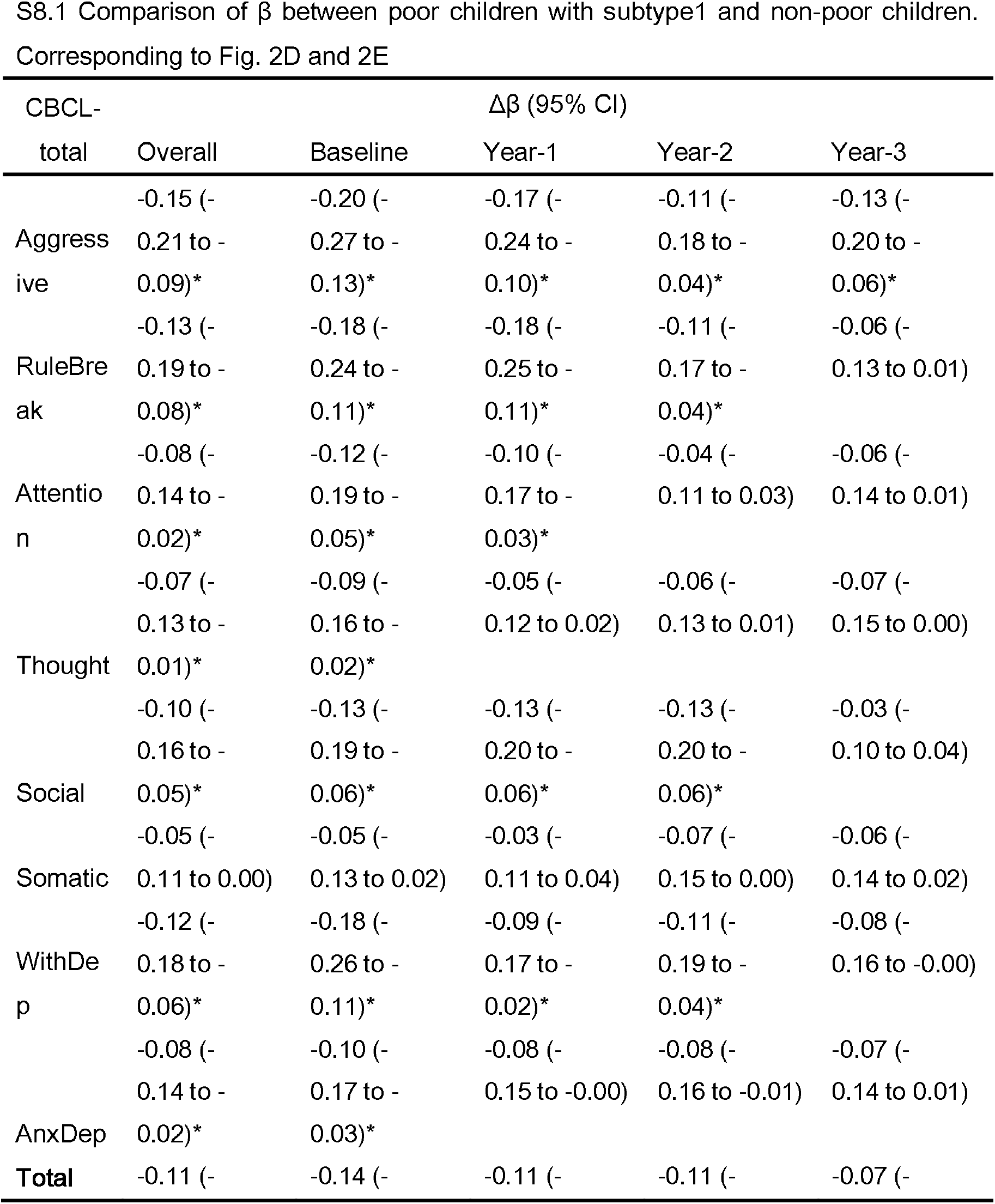
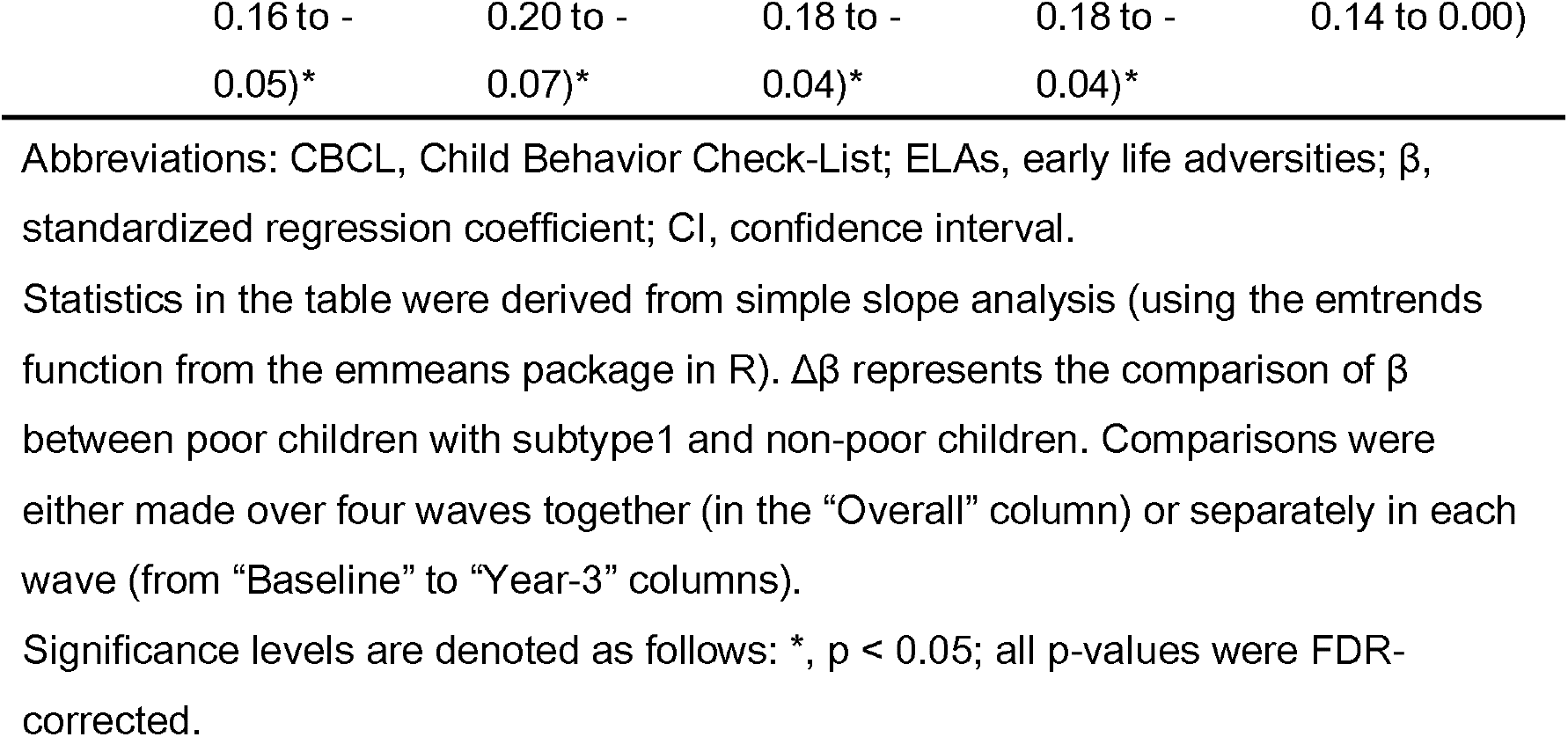
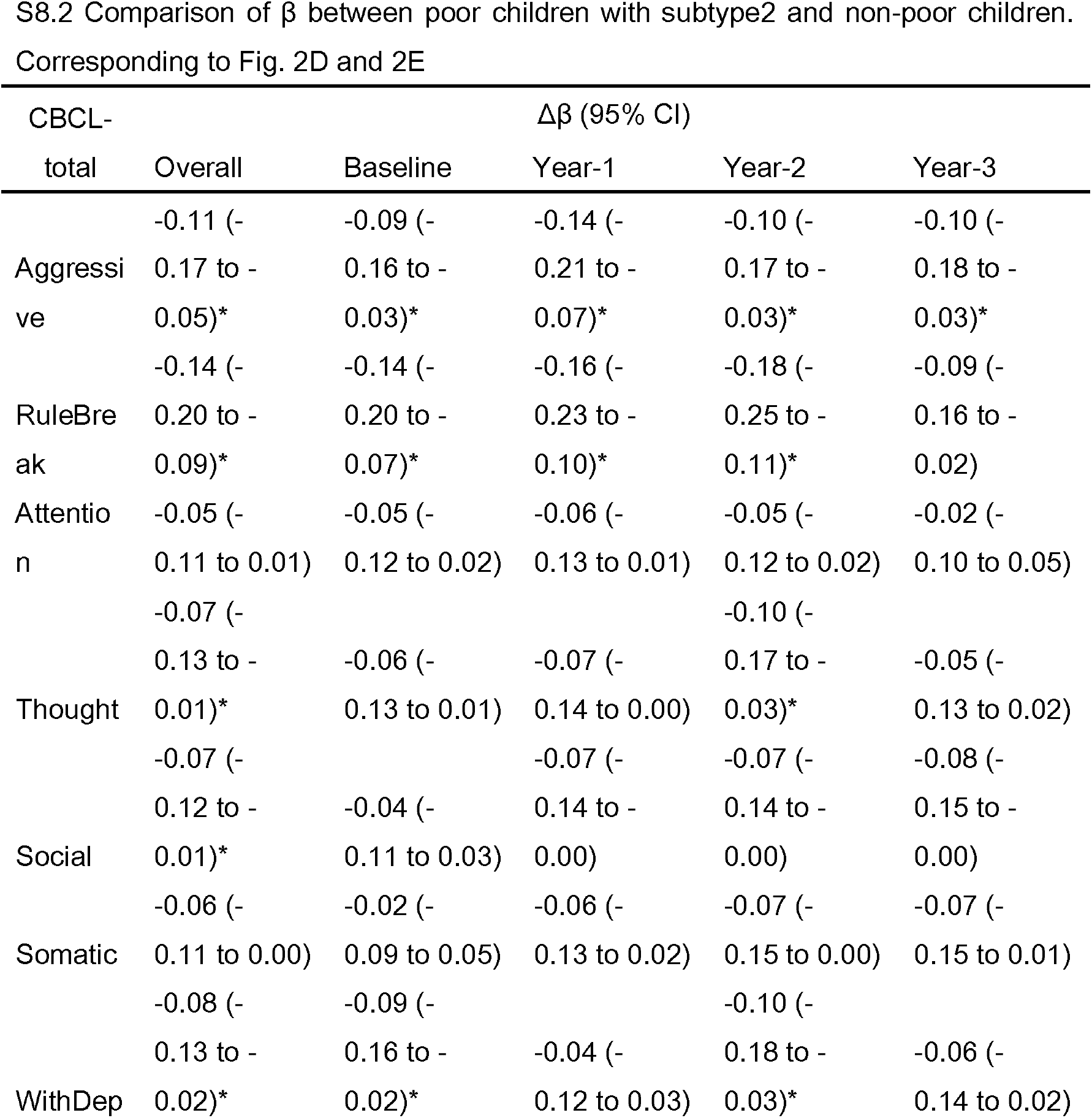
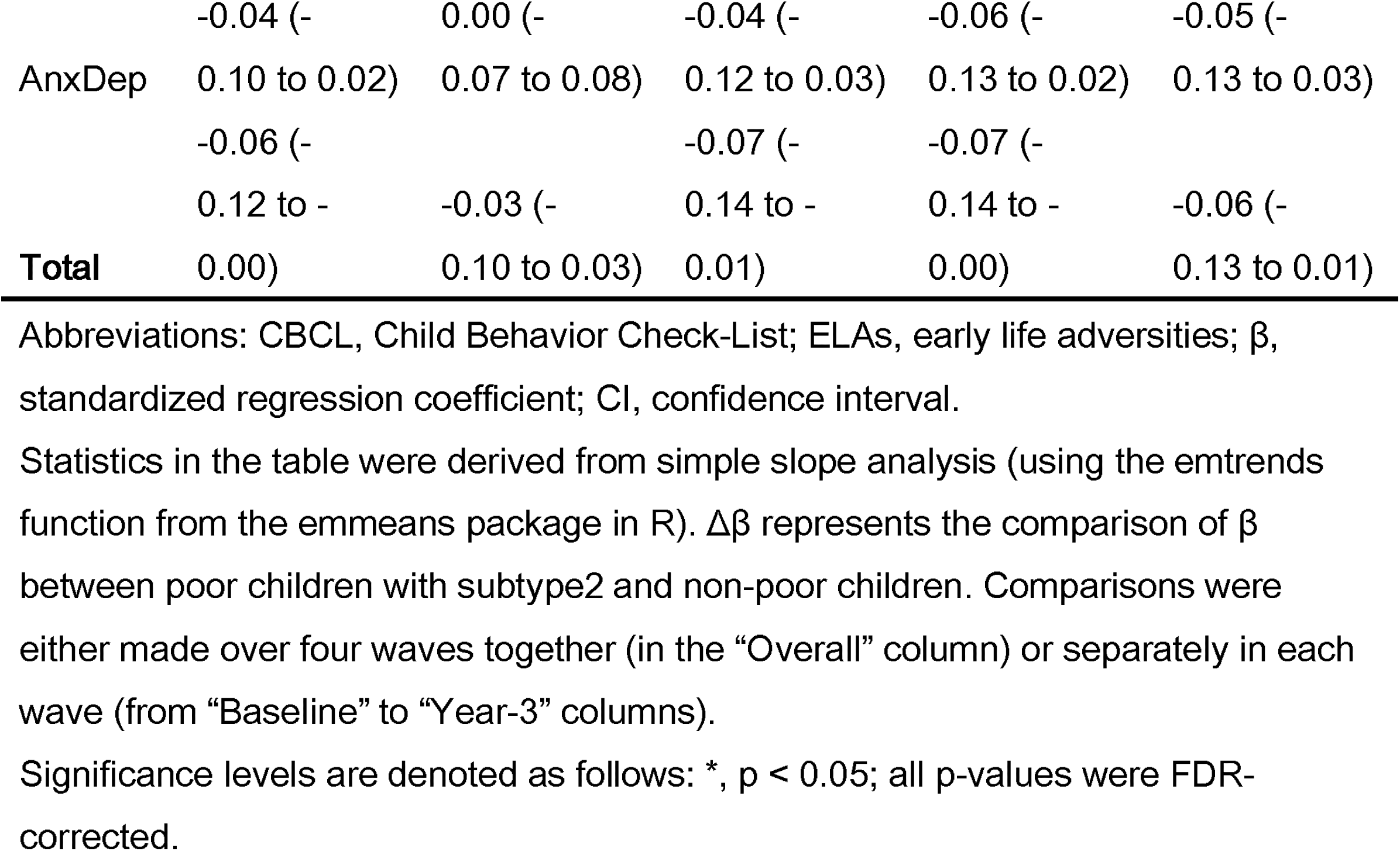

## S9: Difference in superficial characteristics between subtypes

|  | Group summary |  |  | *P* value of<br>subtype1 vs<br>subtype2 |
| --- | --- | --- | --- | --- |
|  | Control | Subtype1 | Subtype2 |  |
| Sex-female, No., (%) | 3212 (49.6) | 235 (48.9) | 225 (51.3) | >.05 |
| Race/Ethnicity (%) | 64/7.5/16.1/2.2/10.2 | 17.7/35.3/34.7/0.8/11.4* | 21.6/31/36.9/0.7/9.8* | >.05 |
| White/Black/Hispanic/Asian/Other |  |  |  |  |
| Age at baseline, mean (SD) | 119.36 (7.55) | 119.16 (7.28) | 118.67 (7.51) | >.05 |
| Log (Income to needs ratio), M<br>(SD) | 5.96 (0.63) | 3.61 (0.82)* | 3.62 (0.79)* | >.05 |
| CBCL-total, M (SD) | 44.97 (10.60) | 46.90 (13.03)* | 46.93 (12.12)* | >.05 |
| Parent has substance abuse<br>problem, No., (%) | 1047 (16.9) | 132 (29.7)* | 122 (29.6)* | >.05 |
| Experience of trauma, No., (%) | 2136 (33.7) | 203 (44.0)* | 187 (43.8)* | >.05 |
| Parental separation, No., (%) | 1090 (16.8) | 296 (62.6)* | 249 (57.2)* | >.05 |
| Experience of being bullied, No.,<br>(%) | 834 (12.9) | 83 (17.3)* | 89 (20.3)* | >.05 |
| Experience of surgery, No., (%) | 1494 (23.0) | 82 (17.0) | 75 (17.1) | >.05 |
| ASR-total, M (SD) | 42.72 (9.62) | 44.40 (12.10)* | 44.21 (11.58)* | >.05 |
| Parent coldness, M (SD)) | 1.21 (0.29) | 1.25 (0.34)* | 1.25 (0.33)* | >.05 |
| Family conflict, M (SD) | 2.45 (1.93) | 2.91 (2.11)* | 2.69 (2.06)* | >.05 |
| Parent neglect, M (SD) | 1.57 (0.46) | 1.69 (0.60)* | 1.72 (0.61)* | >.05 |
| Sleep problem, M (SD) | 0.44 (0.76) | 0.38 (0.71) | 0.38 (0.75) | >.05 |
| Prenatal exposure, M (SD) | 1.73 (1.64) | 2.19 (1.79)* | 2.21 (1.89)* | >.05 |
| Area deprivation index, M (SD) | 34.36 (23.22) | 59.43 (28.70)* | 58.46 (27.40)* | >.05 |
| Neighbourhood insafety, M (SD) | 1.94 (0.86) | 2.70 (1.14)* | 2.66 (1.14)* | >.05 |
| School-bad environment, M (SD) | 10.06 (2.66) | 9.96 (3.16) | 9.91 (3.06) | >.05 |
| Cumulative ELAs-8 sig vars, M | 1.63 (1.05) | 1.92 (1.22)* | 1.92 (1.29)* | >.05 |
| (SD) |  |  |  |  |
| Cumulative ELAs-14 vars, M (SD) | 2.84 (1.36) | 4.05 (1.60)* | 3.99 (1.63)* | >.05 |
| SST-reaction time.i, M (SD) | 285.95 (82.32) | 290.77 (95.68)* | 301.96 (89.67)* | >.05 |
| SST-head motion, M (SD) | 0.39 (0.37) | 0.52 (0.48)* | 0.50 (0.50)* | >.05 |
| Cognition-Picture Vocabulary, M (SD) | 53.30 (11.17) | 50.22 (9.96)* | 50.43 (10.88)* | >.05 |
| Cognition-Flanker, M (SD) | 46.35 (9.04) | 46.42 (9.37) | 46.87 (9.47) | >.05 |
| Cognition-List Sorting, M (SD) | 50.35 (9.64) | 47.76 (10.35)* | 48.05 (10.03)* | >.05 |
| Cognition-Card Sort, M (SD) | 48.36 (9.88) | 46.70 (9.06)* | 46.72 (9.04)* | >.05 |
| Cognition-Pattern Comparison, M (SD) | 46.11 (14.00) | 44.61 (14.88)* | 45.40 (14.56) | >.05 |
| Cognition-Picture Sequence, M (SD) | 50.24 (11.26) | 49.35 (10.38) | 49.01 (10.30) | >.05 |
| Cognition-Oral Reading, M (SD) | 50.67 (11.90) | 45.96 (9.68)* | 46.89 (10.50)* | >.05 |
| Cognition-Fluid Composite, M (SD) | 47.00 (11.08) | 44.85 (11.35)* | 45.21 (10.88)* | >.05 |
| Cognition-Crystallized Composite, M (SD) | 52.35 (11.52) | 47.58 (9.39)* | 48.39 (10.35)* | >.05 |
| Cognition-Total Composite, M (SD) | 49.37 (11.14) | 45.04 (10.42)* | 45.74 (10.60)* | >.05 |

Abbreviations: CBCL, Child Behavior Check-List; ELAs, early life adversities; vars, variables.
Comparisons between groups were made using different statistical models: linear regression model (including sex, age, Income to needs ratio as covariates; only group variable as regressor), logistic regression model (including race/ethnicity as covariate; only group variable as regressor), or mixed-effect models (for all other comparisons; as described in "Comparison of resilience between groups").
Asterisks in the table represent significant comparisons of poor children with different subtypes to non-poor/control children ( $p < .05$ , uncorrected for multiple comparisons).
P values in the last column represent the significance level of the comparison between two subtypes of poor children (since there was no significant result, multiple comparison was not conducted).

## S10: Spatial correlation between neurotransmitter maps and activation maps of 2 subtypes

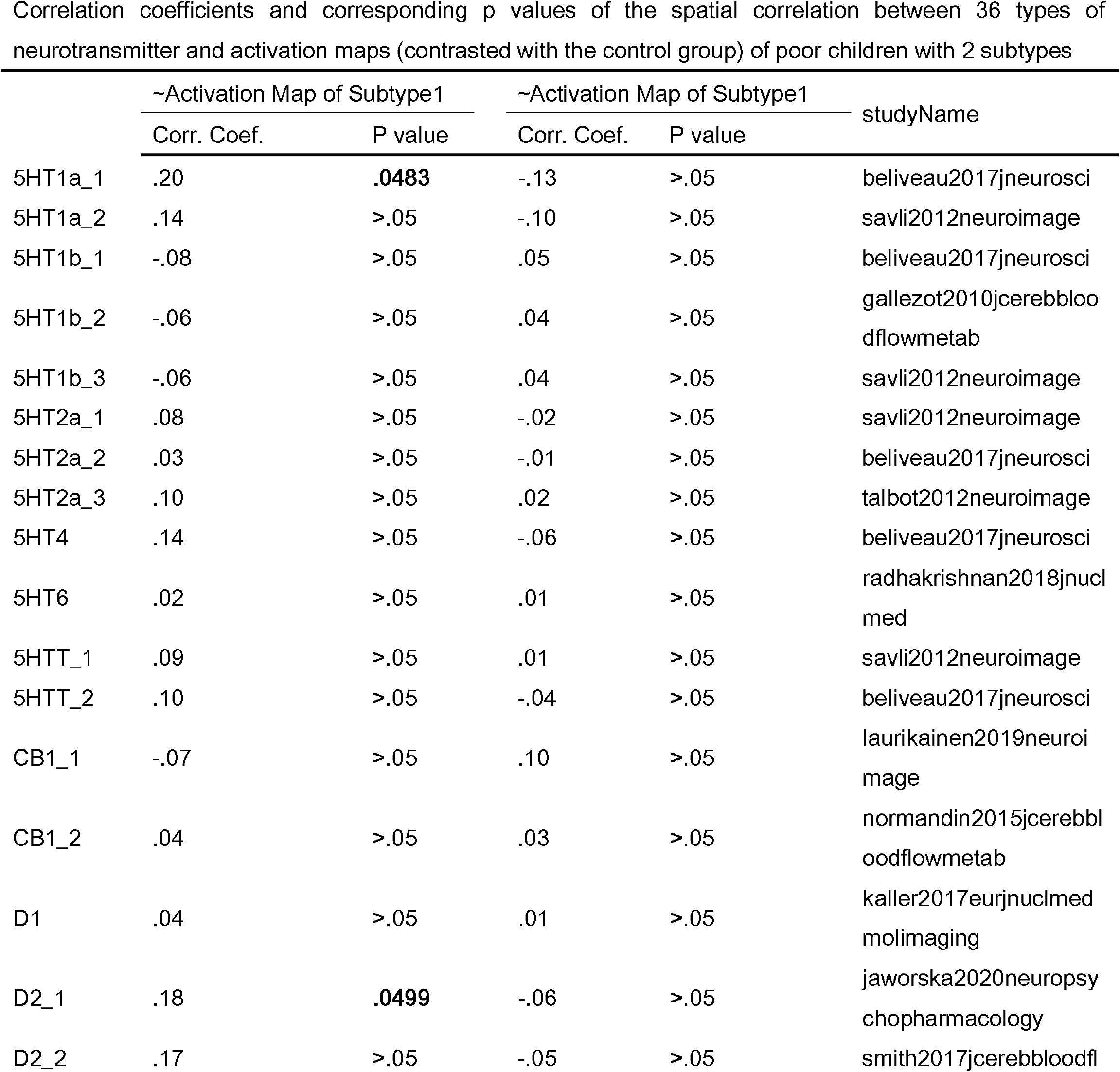
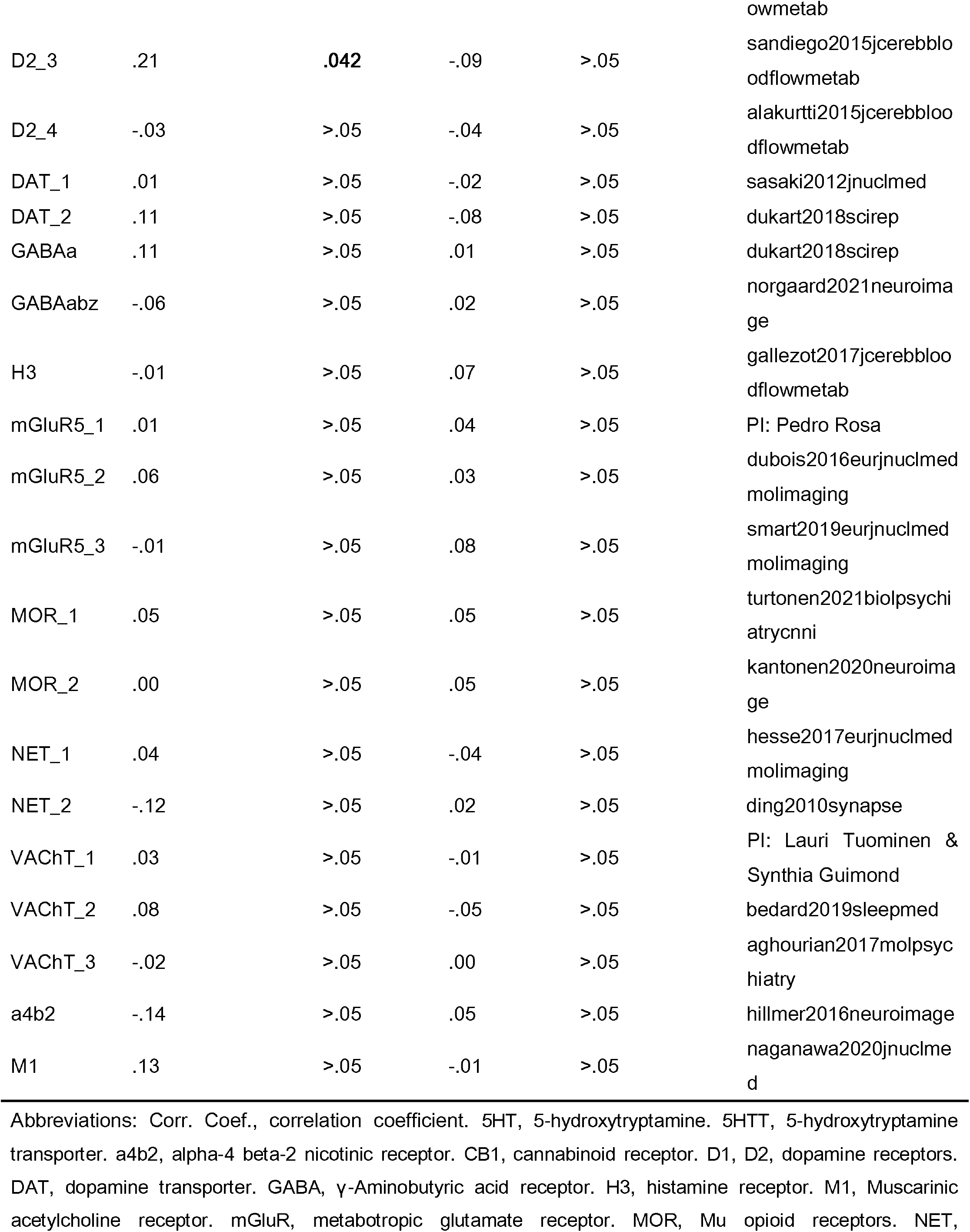
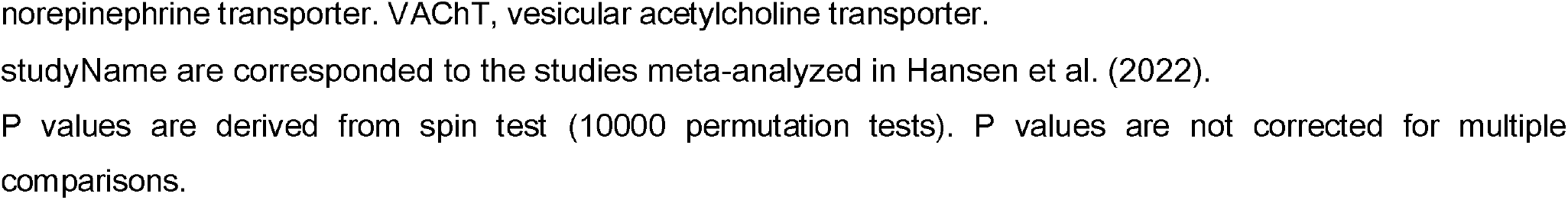

## S11: Sensitivity analysis-1: measuring cumulative ELAs based on all 14 ELAs

In the main analysis, we focused on 8 ELAs that exhibited significant interaction effects with poverty on behavioral problems. These ELAs were chosen to highlight their role in disrupted resilience among poor children, excluding others that did not show such moderation by poverty. However, this approach raises the concern of potentially overlooking the broader impact of ELAs. Therefore, in the first part of sensitivity analysis, we measured cumulative ELAs by considering all 14 ELAs in the study, rather than just the 8 significant ones.

### Poverty × Cumulative ELAs (14 ELAs) -> CBCL

By doing so, we found that compared to their non-poor counterparts, **the impact strength of cumulative ELAs (14 ELAs) on behavior problem was greater in poor children** (β_poor_ (SE) = 0.381 (0.014), β_non-poor_ (SE) = 0.312 (0.004); Δβ (SE) = 0.069 (0.02), *p* < .0001). Then, we further investigated whether the disruption of resilience in poor children persisted into adolescence. Analyzing the impact of cumulative ELAs (14 ELAs) across four waves, we found that childhood poverty continued to amplify this impact. Pooled longitudinal data showed significantly higher impact strength of baseline-measured cumulative ELAs on behavior problems in poor children compared to their non-poor counterparts **over four waves** (β_poor_ (SE) = 0.318 (0.013), β_non-poor_ (SE) = 0.268 (0.007); Δβ (SE) = 0.05 (0.018), *p* = .0004). This impact remained significantly higher even **after 3 years** (β_poor_ (SE) = 0.293 (0.015), β_non-poor_ (SE) = 0.247 (0.008); Δβ (SE) = 0.046 (0.017), *p* = .0073). These results indicate that even when considering all ElAs, rather than only those significantly moderated by poverty, their cumulative effect is still amplified by poverty. This amplification leads to disrupted resilience, and such disrupted resilience can persist into adolescence.

### Subtype × Cumulative ELAs (14 ELAs) -> CBCL

Poor children with **subtype1 showed significantly higher impact strength of baseline-measured cumulative ELAs (14 ELAs) on behavior problems compared to their non-poor counterparts** (β_poor-subtype1_ (SE) = 0.413 (0.025), β_non-poor_ (SE) = 0.299 (0.009); Δβ (SE) = 0.113 (0.026), *p* < .0001), whereas this **difference was not significant in poor children with subtype-2** (β_poor-subtype-2_ (SE) = 0.333 (0.025); Δβ (SE) = 0.034 (0.026), *p* = .201). Even **after 2 years, the impact strength remained significantly higher for poor children with subtype1** (β_poor-subtype1_ (SE) = 0.295 (0.026), β_non-poor_ (SE) = 0.24 (0.009); Δβ (SE) = 0.055 (0.027), *p* = .0413), but not for poor children with subtype-2 (β_poor-subtype-2_ (SE) = 0.283 (0.026); Δβ (SE) = 0.043 (0.028), *p* = .1187). These results indicate that even when considering all ElAs, rather than only those significantly moderated by poverty, resilience is disrupted only for poor children with subtype1 instead of subtype-2.

### L-MFG × Cumulative ELAs (14 ELAs) -> CBCL

We then examined whether L-MFG thickness provided resilience against the cumulative ELAs (14 ELAs) in poor children with subtype1. Compared to children with the lowest L-MFG thickness, those with the highest L-MFG thickness demonstrated significantly lower impact strength of cumulative ELAs on behavior problems (β_high_ (SE) = 0.285 (0.043), β_low_ (SE) = 0.406 (0.042); Δβ (SE) = 0.12 (0.059), *p* = .0419). Even after 3 years, the impact strength remained significantly lower for individuals with the highest thickness (β_high-thick_ (SE) = 0.162 (0.048), β_low-thick_ (SE) = 0.314 (0.047); Δβ (SE) = 0.152 (0.066), *p* = .0216). These findings highlight the resilient role of L-MFG against cumulative ELAs exclusively in poor children with subtype1. These results indicate that even when considering all ELAs, rather than only those significantly moderated by poverty, their cumulative effect is still modulated by the L-MFG thickness. Specifically, greater L-MFG thickness can provide resilience effect in poor children with subtype1.

## S12: Sensitivity analysis-2: measuring cumulative ELAs based on complete cases

In our primary analysis, we addressed missing data in cumulative ELAs by employing mean value imputation for each ELA type. However, recognizing the potential confounding effects of this imputation method, we conducted a secondary analysis. Specifically, we recalculated cumulative ELAs using only participants with complete data across all eight ELA categories. Among the initial 10,112 children in the study, we identified missing data as follows: 490 (4.8%) for ’Parental Substance Abuse,’ 266 (2.6%) for ’Experience of Trauma,’ 2 (0.02%) for ’Experience of Being Bullied,’ 1 (0.01%) for ’Experience of Surgery,’ 54 (0.53%) for ’ASR-total,’ 0 for ’Parental Coldness,’ 202 (2.0%) for ’Sleep Problems,’ and 1,432 (14.2%) for ’Prenatal Exposure.’ Following the exclusion of these cases from the dataset, we retained 8094 children with complete ELA measures for further analysis.

### Poverty × Cumulative ELAs (8 ELAs with complete cases) -> CBCL

By doing so, we found that compared to their non-poor counterparts, **the impact strength of cumulative ELAs (8 ELAs with complete cases) on behavior problem was greater in poor children** (β_poor_ (SE) = 0.481 (0.019), β_non-poor_ (SE) = 0.387 (0.01); Δβ (SE) = 0.094 (0.022), *p* < .0001). Then, we further investigated whether the disruption of resilience in poor children persisted into adolescence. Analyzing the impact of cumulative ELAs (8 ELAs with complete cases) across four waves, we found that childhood poverty continued to amplify this impact. Pooled longitudinal data showed significantly higher impact strength of baseline-measured cumulative ELAs on behavior problems in poor children compared to their non-poor counterparts **over four waves** (β_poor_ (SE) = 0.408 (0.017), β_non-poor_ (SE) = 0.326 (0.009); Δβ (SE) = 0.083 (0.02), *p* < .0001). This impact remained significantly higher even **after 3 years** (β_poor_ (SE) = 0.376 (0.021), β_non-poor_ (SE) = 0.302 (0.011); Δβ (SE) = 0.074 (0.024), *p* = .0019). These results indicate that the poverty’s disruption of resilience in the main analysis cannot be attributed to the handling of missing values.

### Subtype × Cumulative ELAs (8 ELAs with complete cases) -> CBCL

Poor children with **subtype1 showed significantly higher impact strength of baseline-measured cumulative ELAs (8 ELAs with complete cases) on behavior problems compared to their non-poor counterparts** (β_poor-subtype1_ (SE) = 0.512 (0.034), β_non-poor_ (SE) = 0.377 (0.011); Δβ (SE) = 0.135 (0.036), *p* = .0002), whereas this **difference was not significant in poor children with subtype-2** (β_poor-subtype-2_ (SE) = 0.428 (0.035); Δβ (SE) = 0.083 (0.048), *p* = .081). Even **after 2 years, the impact strength remained significantly higher for poor children with subtype1** (β_poor-subtype1_ (SE) = 0.409 (0.037), β_non-poor_ (SE) = 0.294 (0.012); Δβ (SE) = 0.115 (0.038), *p* = .0028), but not for poor children with subtype-2 (β_poor-subtype-2_ (SE) = 0.363 (0.037); Δβ (SE) = 0.069 (0.039), *p* = .0753). These results indicate that the subtype2’s disruption of resilience for poor children in the main analysis cannot be attributed to the handling of missing values.

### L-MFG × Cumulative ELAs (8 ELAs with complete cases) -> CBCL

We then examined whether L-MFG thickness provided resilience against the cumulative ELAs (8 ELAs with complete cases) in poor children with subtype1. Compared to children with the lowest L-MFG thickness, those with the highest L-MFG thickness demonstrated lower impact strength of cumulative ELAs on behavior problems which was **only marginally significant** (β_high_ (SE) = 0.509 (0.063), β_low_ (SE) = 0.368 (0.056); Δβ (SE) = 0.14 (0.084), *p* = .097). However, **pooled longitudinal data showed significantly lower impact strength of baseline-measured cumulative ELAs** (8 ELAs with complete cases) on behavior problems for individuals with the highest thickness (β_high-thick_ (SE) = 0.234 (0.053), β_low-thick_ (SE) = 0.44 (0.059); Δβ (SE) = 0.206 (0.079), *p* = .0091) **over four waves**. Even **after 3 years**, the impact strength remained significantly lower for individuals with the highest thickness (β_high-thick_ (SE) = 0.138 (0.067), β_low-thick_ (SE) = 0.35 (0.075); Δβ (SE) = 0.211 (0.1), *p* = .0348). These results indicate that the resilience effect of L-MFG thickness in poor children with subtype1 cannot be attributed to the handling of missing values.

## S13: Sensitivity analysis-3: measuring cumulative ELAs based on all 14 ELAs

Oversampling for siblings and twins is a notable characteristic of the ABCD study (Saragosa-Harris et al., 2022). Out of the 10,112 children included in our analysis, 1,700 (16.8%) have siblings who are also participants in the study. This introduces a family-level confounder, namely twin- or sibling-effects, into our data analysis, which has been shown to significantly influence mental health outcomes in the context of Early Life Adversities (ELAs) (Daníelsdóttir et al., 2024). While the main analysis controlled for family-level confounders by employing modeling with a nested structure (three-level models, children/family/site; Saragosa-Harris et al., 2022), we conducted an additional analysis to thoroughly exclude family-level confounding. This involved randomly selecting one sibling from each sibling pair, resulting in a dataset comprising 8414 children.

### Poverty × Cumulative ELAs -> CBCL

By doing so, we found that compared to their non-poor counterparts, **the impact strength of cumulative ELAs (8 ELAs with significant moderated effect) on behavior problem was greater in poor children** (β_poor_ (SE) = 0.497 (0.019), β_non-poor_ (SE) = 0.41 (0.01); Δβ (SE) = 0.087 (0.021), *p* < .0001). Then, we further investigated whether the disruption of resilience in poor children persisted into adolescence. Analyzing the impact of cumulative ELAs across four waves, we found that childhood poverty continued to amplify this impact. Pooled longitudinal data showed significantly higher impact strength of baseline-measured cumulative ELAs on behavior problems in poor children compared to their non-poor counterparts **over four waves** (β_poor_ (SE) = 0.431 (0.017), β_non-poor_ (SE) = 0.358 (0.009); Δβ (SE) = 0.073 (0.019), *p* = .0001). This impact remained significantly higher even **after 3 years** (β_poor_ (SE) = 0.404 (0.021), β_non-poor_ (SE) = 0.335 (0.01); Δβ (SE) = 0.068 (0.023), *p* = .0034). These results indicate that even when considering all ElAs, rather than only those significantly moderated by poverty, their cumulative effect is still amplified by poverty. These results indicate that the poverty’s disruption of resilience in the main analysis cannot be attributed to the family-level confounders.

### Subtype × Cumulative ELAs -> CBCL

Poor children with **subtype1 showed significantly higher impact strength of baseline-measured cumulative ELAs (8 ELAs with significant moderated effect) on behavior problems compared to their non-poor counterparts** (β_poor-subtype1_ (SE) = 0.548 (0.034), β_non-poor_ (SE) = 0.391 (0.011); Δβ (SE) = 0.157 (0.036), *p* < .0001), whereas this **difference was not significant in poor children with subtype-2** (β_poor-subtype-2_ (SE) = 0.452 (0.036); Δβ (SE) = 0.061 (0.038), *p* = .108). Even **after 2 years, the impact strength remained significantly higher for poor children with subtype1** (β_poor-subtype1_ (SE) = 0.422 (0.036), β_non-poor_ (SE) = 0.316 (0.012); Δβ (SE) = 0.106 (0.038), *p* = .0055), but not for poor children with subtype-2 (β_poor-subtype-2_ (SE) = 0.393 (0.039); Δβ (SE) = 0.077 (0.041), *p* = .0578). These results indicate that the subtype2’s disruption of resilience for poor children in the main analysis cannot be attributed to the family-level confounders.

### L-MFG × Cumulative ELAs -> CBCL

We then examined whether L-MFG thickness provided resilience against the cumulative ELAs (8 ELAs with significant moderated effect) in poor children with subtype1. Compared to children with the lowest L-MFG thickness, those with the highest L-MFG thickness demonstrated lower impact strength of cumulative ELAs on behavior problems which was **only marginally significant** (β_high_ (SE) = 0.431 (0.058), β_low_ (SE) = 0.566 (0.061); Δβ (SE) = 0.135 (0.082), *p* = .1025). However, **pooled longitudinal data showed significantly lower impact strength of baseline-measured cumulative ELAs** on behavior problems for individuals with the highest thickness (β_high-thick_ (SE) = 0.353 (0.053), β_low-thick_ (SE) = 0.502 (0.056); Δβ (SE) = 0.15 (0.076), *p* = .048) **over four waves**. These results indicate that the resilience effect of L-MFG thickness in poor children with subtype1 cannot be fully attributed to the family-level confounders.

